# Plasma biomarker levels and cognitive decline in a heterogenous community-based cohort with multiple comorbidities

**DOI:** 10.64898/2026.05.29.26354375

**Authors:** Marc D. Rudolph, Julia R. Bacci, Jillian K. Lee, Sarah A. Gaussoin, James R. Bateman, Timothy M. Hughes, Shannon L. Risacher, Laura D. Baker, Goldie S. Byrd, Courtney L. Sutphen, Thomas C. Register, Michelle M. Mielke, Suzanne Craft

**Author notes:** **Corresponding author:** Marc D. Rudolph, PhD, Wake Forest School of Medicine, Medical Center Blvd, Winston-Salem, NC 27157.

## Abstract

**INTRODUCTION:** Knowledge about how Alzheimer’s disease (AD) and AD-related dementia (AD/ADRD) plasma biomarkers relate to global and domain-specific cognitive functioning across diagnostic groups remains limited, particularly in heterogeneous, community-dwelling populations with multiple comorbidities.

**METHODS:** We evaluated associations between baseline plasma biomarker levels (Aβ42/40, p-tau181, p-tau217, NfL, GFAP) and cognitive performance at baseline and longitudinally (up to 7 years). Participants (n=590) enrolled in the Wake Forest Alzheimer’s Disease Research Center Clinical Core (314 cognitively unimpaired [CU]; 206 mild cognitive impairment [MCI]; and 70 dementia) completed annual cognitive assessments including the Uniform Data Set (UDSv3; NACC). Domain-specific cognitive composites including memory, executive function, attention, language, visuospatial ability, and phonemic fluency, as well as a modified Preclinical Alzheimer’s Cognitive Composite (PACC5), were evaluated. General linear and mixed-effects models were adjusted for demographics (age, sex, race, education), APOE-ε4 status, comorbidities (estimated glomerular filtration rate; BMI), and cardiometabolic health factors (hypertension, diabetes). Effect modification by cognitive diagnosis was evaluated.

**RESULTS:** Baseline plasma biomarkers, particularly p-tau217, were associated with poorer baseline cognitive performance and greater longitudinal decline on the PACC5 and all cognitive domains assessed, except phonemic fluency (strongest for memory). Post-hoc analyses indicated associations between plasma biomarker levels and cognition were generally more pronounced in MCI compared with CU participants. Effect modification by baseline cognitive status was limited and attenuated when all biomarkers were modeled simultaneously. Comorbidities and cardiometabolic factors modified select associations.

**DISCUSSION:** Plasma AD/ADRD biomarkers, particularly p-tau217, were associated with cognitive impairment and decline in a heterogenous community cohort.

**HIGHLIGHTS:**

- The WF ADRC is a clinically heterogeneous, comorbidity-rich community-dwelling cohort
- Plasma AD/ADRD biomarkers were associated with poorer baseline cognition and longitudinal cognitive decline
- Associations were strongest for the PACC5 composite and memory-related cognitive domains
- p-tau217 provided the most robust biomarker signal across models

## 1 INTRODUCTION

Plasma biomarkers have emerged as minimally invasive and cost-effective measures for detecting Alzheimer’s disease (AD) pathology.^1–5^ Key plasma biomarkers for AD and related dementias (ADRD) include the ratio of amyloid beta 1-42 (Aβ42) to 1-40 (Aβ40), phosphorylated tau 181 and 217 (p-tau181; p-tau217), neurofilament-light chain (NfL), and glial fibrillary acidic protein (GFAP). These biomarkers have been associated with poorer overall brain health,^1,6–11^ worse daily functioning,^12–14^ and impaired cognitive abilities.^12,15–31^ As previously described, plasma-derived analytes of amyloid (Aβ40, Aβ42) and phosphorylated tau (p-tau181; p-tau217) are AD-specific peripheral biomarkers sensitive to brain amyloid plaque deposition,^5,11,32–42^ whereas NfL and GFAP are non-specific markers of axonal neurodegeneration and neuroinflammation, respectively, that can contribute to cognitive impairment and mixed pathological forms of ADRD.^3,5,24,43^

Prior studies have primarily examined associations between a limited subset of plasma biomarkers and either single cognitive domain scores or global cognitive screening instruments.^19,25,28,44,45^ While these studies have demonstrated associations between plasma biomarkers and cognitive performance, it remains unclear whether these relationships persist after adjusting for potential confounders, including common comorbidities and cardiometabolic factors.^18,33^ Moreover, most prior work has been conducted in highly selected research cohorts where medical comorbidities are not consistently collected or well characterized, limiting generalizability to the community populations in which scalable biomarkers are most needed. Importantly, few studies have evaluated domain-specific cognitive outcomes, despite evidence that subtle changes in episodic memory and executive function can emerge earlier than detectable alterations on global cognitive measures.^46–49^ Finally, comorbidities such as chronic kidney disease, obesity, hypertension, and diabetes may influence both biomarker concentrations and cognitive performance, potentially confounding or modifying biomarker–cognition associations.^18,33^ Thus, further work is needed to evaluate the cross-sectional and longitudinal relationships between plasma biomarkers and cognition in community-representative, heterogeneous cohorts with multiple comorbidities.

In the present study, we comprehensively examined the cross-sectional and longitudinal associations between baseline AD/ADRD plasma biomarker levels (Aβ42/Aβ40, p-tau181, p-tau217, NfL, GFAP) and measures of global and domain-specific cognition among a cohort of participants with up to 7 years of follow-up. We leveraged data from the Wake Forest Alzheimer’s Disease Research Center (WFADRC), a heterogeneous community-dwelling cohort located in the piedmont/triad area of western North Carolina and the surrounding region with a high incidence of comorbidities and chronic conditions associated with elevated cardiometabolic risk. We hypothesized that lower levels of Aβ42/Aβ40 and higher levels of p-tau181, p-tau217, NfL, or GFAP would be associated with worse cognition cross-sectionally and greater cognitive change from baseline longitudinally. Among plasma biomarkers, p-tau217 has demonstrated the highest accuracy for detecting AD pathology, and has shown greater sensitivity to early cognitive change than p-tau181, NfL, Aβ42/40, or GFAP in multiple cohorts, including recent work from our group and others.^25,37,38,50–53^ Given that episodic memory is among the earliest and most prominent cognitive deficits observed in typical AD and is heavily represented in the Uniform Data Set (UDS) and in memory-weighted composites such as the PACC5,^54–56^ we hypothesized that p-tau217 would show the strongest associations with cognition, particularly in the memory domain. We further hypothesized that these associations would be most pronounced among participants with mild cognitive impairment (MCI), who may be in a transitional stage along the ADRD continuum where biomarker–cognition relationships are most detectable.

## 2 METHODS & MATERIALS

### 2.1 Participants

All participants were enrolled in the WFADRC, a representative community-based cohort that focuses on the transition from normal aging to MCI and dementia, with specific emphasis on understanding the contribution of metabolic and vascular factors to these transitions. Adults between the ages of 55 and 85 were recruited from the surrounding community via the WFADRC into the Clinical Core supported by efforts of the Outreach, Engagement & Recruitment Core.^1,57^ As described previously, this was a community sample based in North Carolina and surrounding areas that have higher rates of obesity, hypertension, cardiovascular and other disease compared to the US average.^1,57,58^ Participants underwent a standard evaluation, including the National Alzheimer’s Coordinating Center (NACC) protocol for clinical research data collection, clinical exams, neurocognitive testing, neuroimaging, and genotyping for the apolipoprotein E (*APOE*) ε4 allele. Exclusion criteria included a history of large vessel stroke (participants with lacunae or small vessel ischemic disease were eligible); other significant neurologic diseases; uncontrolled *chronic* medical or psychiatric conditions (such as advanced liver or severe kidney disease [estimated glomerular filtration rate; eGFR<30]; poorly controlled congestive heart failure, chronic obstructive pulmonary disease or sleep apnea; active cancer treatment; uncontrolled clinical depression, or psychiatric illness; current use of insulin; and history of substance abuse or heavy alcohol consumption within the previous ten years). The Wake Forest Institutional Review Board approved all activities as described; written informed consent was obtained for all participants and/or their legally authorized representatives according to the Declaration of Helsinki.

### 2.2 Assessment of Demographics and Health Conditions

Participants self-reported age, race/ethnicity (e.g., Black/White; Non-Hispanic), sex (e.g., male/female), and years of education. *APOE* genotype was obtained by Taqman using single nucleotide polymorphisms (rs429358 and rs7412) to determine haplotypes of ε2, ε3, and ε4. *APOE* was dichotomized to represent the presence or absence of one or more ε4 alleles (e.g., carrier vs. non-carrier; *APOE*-*ε*4). BMI was calculated as weight (lb)/[height (in)]^2^ x 703. Kidney function was ascertained using eGFR values from LabCorp assay calculated with the updated, race-free CKD-EPI 2021 equation (https://www.labcorp.com/tests/100768/glomerular-filtration-rate-estimated-egfr). Fasting blood glucose levels were measured via an oral glucose tolerance test (OGTT) from serial blood draws; impaired glucose tolerance was defined by glucose ≥ 140 mg/dL at 120 minutes of OGTT or hemoglobin A1c (HbA1c) ≥ 5.7% as previously described.^59^ Diabetic status (normal; prediabetic; diabetic) was determined using a combination of OGTT data and hemoglobin A1c (HbA1c) levels. Participants were coded as diabetic if HbA1C ≥ 6.5%, OGTT (time 0) ≥ 126 mg/dL, or OGTT (120 minutes) ≥ 200mg/dL, or as prediabetic if HbA1c was between 5.7% and 6.5% or OGTT (time 0) was between 100 and 126 mg/dL or OGTT (120 minutes) was between 140 and 200 mg/dL. Hypertension status (HTN) was defined according to 2017 ACC/AHA guidelines as seated blood pressure ≥ 130 mmHg, diastolic blood pressure ≥ 80 mmHg, and/or current use of antihypertensive medications as described elsewhere.^1^

### 2.3. Cognitive Testing

Participants completed cognitive testing with the UDS Version 3 (UDSv3) neuropsychological battery which included: Montreal Cognitive Assessment (MoCA), Craft Story, Benson Figure, Number Span, Phonemic Fluency (letters F and L), Category Fluency (Animals and Vegetables), Trail Making Test A and B, and the Multilingual Naming Test.^54^ Participants also completed the Mini‐Mental State Examination (MMSE) and additional assessments capturing memory functioning (Free and Cued Selective Reminding Test [FCSRT], Rey Auditory Verbal Learning Test [AVLT]) and executive functioning and processing speed (Digit Symbol Substitution Test [DSST]). UDSv3 cognitive assessments were adjusted for age, self‐reported race, sex, and education and converted to domain specific cognitive performance Z‐scores for: executive function, memory, language, attention, visuospatial, and phonemic domains,^54,60^ as described previously.^57^ A modified Preclinical Alzheimer’s Cognitive Composite (PACC5) was generated using scores from the MMSE, AVLT delayed-recall, Craft Story delayed verbatim recall, DSST, and Category Fluency assessments.^48,49^

### 2.4 Diagnostic Adjudication

An expert panel of investigators, including neuropsychologists, neurologists, and geriatricians, provided adjudication of cognitive status as either cognitively unimpaired (CU), MCI,^61^ or dementia (DEM).^62^ Adjudication of cause/type of cognitive impairment or dementia was assessed using neuroimaging and fluid biomarkers.

### 2.5 Plasma Biomarkers

Plasma AD/ADRD biomarkers were collected from subjects after a minimum 8-hour (water only) fast. Blood was processed within 30 minutes of collection, and plasma biomarkers were assessed as previously described.^1^ Briefly, batch shipments of frozen aliquots were sent to the National Centralized Repository for Alzheimer’s Disease and Related Dementias (NCRAD) Biomarker Assay Laboratory on dry ice for analysis of plasma Aβ42, Aβ40, NfL, and GFAP using the Quanterix Simoa Neurology 4-Plex E and plasma p-tau181 v2 Advantage Kits on a Quanterix Simoa HD-X. Plasma p-tau217 was assessed using ALZpath Simoa p-tau 217 v2 assay kits on a Quanterix HD-X at Neurocode (Bellingham WA). Additional details can be found in Rudolph et al. (2025).^1^

### 2.6 Statistical Analysis

Unadjusted associations between baseline plasma biomarker levels, cognitive performance, and health-related factors were evaluated using Spearman’s rank correlation, which is appropriate for non-parametric and potentially non-linear relationships and consistent with prior work.^16^ Associations between baseline plasma biomarker levels and baseline cognition were examined using general linear models (GLMs); linear mixed effects models were used to evaluate associations between baseline plasma biomarker levels and longitudinal change in cognition over time. GLMs were estimated using robust standard errors. Linear mixed effects models included participant-level random intercepts to account for repeated cognitive assessments. Independent multivariable models, in which each biomarker was entered separately along with covariates, were used to estimate the total association between *each* biomarker and cognition. Simultaneous multivariable models, in which all biomarkers were entered *together* along with covariates, were used to assess biomarker-specific associations after accounting for shared variance among correlated biomarkers. This complementary modeling approach is appropriate when biomarkers reflect overlapping but biologically distinct pathological processes.^63,64^ Independent and simultaneous multivariable regression models were adjusted for baseline demographic, genetic, and health-related factors. Model 1 included age, sex, race, ethnicity, education, and APOE-ε4 carriership. Model 2 additionally adjusted for BMI, eGFR, diabetes, and hypertension. Longitudinal models further adjusted for time between cognitive assessments. Plasma biomarkers were log-transformed (except Aβ42/40) and subsequently standardized (centered and scaled by their standard deviation) prior to analysis to facilitate comparison of effect sizes across biomarkers.^16^ Multiple-comparison correction was performed using the Benjamini–Hochberg false discovery rate (FDR) procedure, as applicable.^65^ Collinearity among predictors was assessed using variance inflation factors (VIFs), with values <5 considered acceptable; more conservative thresholds (VIF<2.5) were also considered when interpreting models.^66^ All VIFs were within acceptable limits (range: 1.00–2.82). VIFs for p-tau217 ranged from 2.10–2.82, reflecting moderate correlation with p-tau181; exclusion of p-tau181 did not materially alter results or interpretation and therefore is not discussed further. Post-hoc analyses examined whether baseline cognitive status modified associations between plasma biomarkers and longitudinal cognitive change among participants adjudicated as CU or MCI. Participants adjudicated with dementia were excluded from these analyses due to limited longitudinal follow-up. Cognitive conversion from baseline diagnosis was assessed by comparing participants who progressed from their baseline status to MCI or dementia during follow-up (converters) with those who had the same cognitive status at baseline and at their final assessment (stable). In sensitivity analyses, inverse probability weighting was used in linear mixed effects models to evaluate the potential impact of differential attrition on longitudinal findings. Results were qualitatively similar to primary analyses and are not discussed further. Descriptive analyses, GLMs, and mixed effects models were conducted using R (RStudio Team, 2020) and SAS/STAT software, version 9.4 (SAS Institute Inc., Cary, NC).

## 3 RESULTS

Participant characteristics are shown in Table 1. The mean (SD) age of participants was 70.0 (8.01) years; ∼19% self-identified as Black/African American, 65% as female, and 34% were *APOE*-ε4 carriers. Participants in the WFADRC with baseline plasma biomarker data had up to 7 years of additional follow-up cognitive data; approximately 70% of the cohort had 3 or more visits with an average of 1,221 days (3.3 years) between the first and last cognitive assessment (see Figure S1). About 10% (*n*=60) of participants progressed to MCI or DEM during the study (Table S1). Compared to those who remained stable in their baseline diagnosis, progressors, on average, were (1) older; (2) had lower eGFR; (3) had higher plasma biomarker levels and a lower Aβ42/40 ratio; and (3) had poorer cognition across cognitive assessments (Table S1). Baseline cognitive outcomes were significantly correlated (*rho*=.09-.71; Figure S2). PACC5 and memory domain z-scores exhibited the strongest correlations (*rho*=.71; *p*<.001); the visuospatial domain was weakly correlated with all other cognitive domains (memory, executive, language, attention, and phonemic; *rho* =.09-.22; Figure S2).

**Table 1.** Baseline participant characteristics.

| | Overall<br>N = 590<sup>1</sup> | CU<br>N = 314<sup>1</sup> | MCI<br>N = 206<sup>1</sup> | DEM<br>N = 70<sup>1</sup> | $p^2$ |
| --- | --- | --- | --- | --- | --- |
| Age (Years) | 70 (8) | 69 (8) | 72 (8) | 73 (8) | <.001 |
| Female | 382 (65%) | 227 (72%) | 119 (58%) | 36 (51%) | <.001 |
| Race |  |  |  |  | .023 |
| White | 473 (80%) | 257 (82%) | 156 (76%) | 60 (86%) |  |
| AI/AN | 2 (0.3%) | 0 (0%) | 1 (0.5%) | 1 (1.4%) |  |
| Asian | 5 (0.8%) | 5 (1.6%) | 0 (0%) | 0 (0%) |  |
| Black/AA | 110 (19%) | 52 (17%) | 49 (24%) | 9 (13%) |  |
| Hispanic (Yes) | 11 (1.9%) | 5 (1.6%) | 5 (2.4%) | 1 (1.4%) | .800 |
| Education (Years) | 15.77 (2.51) | 16.13 (2.32) | 15.26 (2.64) | 15.67 (2.74) | <.001 |
| *APOE-ε4* (Carrier) | 193 (34%) | 83 (27%) | 72 (36%) | 38 (57%) | <.001 |
| eGFR | 79 (15) | 79 (14) | 78 (16) | 77 (14) | .500 |
| BMI | 0.92 (0.12) | 0.90 (0.12) | 0.93 (0.11) | 0.94 (0.12) | .001 |
| Diabetes |  |  |  |  | .130 |
| Normal | 211 (36%) | 113 (36%) | 67 (33%) | 31 (44%) |  |
| Prediabetes | 298 (51%) | 163 (53%) | 103 (50%) | 32 (46%) |  |
| Diabetes | 77 (13%) | 34 (11%) | 36 (17%) | 7 (10%) |  |
| Hypertension | 257 (44%) | 113 (36%) | 110 (53%) | 34 (49%) | <.001 |
| Aβ42/40 | 0.053 (0.011) | 0.055 (0.010) | 0.051 (0.011) | 0.049 (0.007) | <.001 |
| p-tau181 | 3.46 (2.00) | 2.98 (1.66) | 3.66 (2.18) | 5.07 (1.96) | <.001 |
| p-tau217 | 0.43 (0.36) | 0.32 (0.22) | 0.49 (0.39) | 0.87 (0.51) | <.001 |
| NfL | 17 (14) | 15 (8) | 19 (20) | 24 (11) | <.001 |
| GFAP | 137 (76) | 119 (59) | 142 (83) | 200 (84) | <.001 |
| PACC5 | -0.71 (1.13) | 0.00 (0.64) | -1.22 (0.71) | -2.64 (0.97) | <.001 |
| Memory Domain | -0.47 (1.16) | 0.21 (0.67) | -0.84 (0.80) | -2.43 (1.06) | <.001 |
| Language Domain | -0.32 (0.89) | 0.08 (0.60) | -0.53 (0.69) | -1.53 (1.18) | <.001 |
| Attention Domain | -0.38 (1.09) | 0.00 (0.64) | -0.50 (0.84) | -1.72 (1.90) | <.001 |
| Executive Domain | -0.40 (0.98) | 0.07 (0.64) | -0.71 (0.89) | -1.57 (1.12) | <.001 |
| Visuospatial Domain | -0.35 (1.42) | -0.03 (0.78) | -0.43 (1.09) | -1.56 (3.01) | <.001 |
| Phonemic Domain | -0.30 (0.99) | 0.02 (0.90) | -0.56 (0.90) | -0.98 (1.07) | <.001 |

<sup>1</sup> n (%); Mean (SD); <sup>2</sup> Pearson's Chi-squared test; Kruskal-Wallis rank sum test; Fisher's exact test
**Note:** Baseline participant characteristics are presented. **Abbreviations:** CU = Cognitively Normal; MCI = Mild Cognitive Impairment; DEM = Dementia; AI/AN = American Indian or Alaskan Native; AA = African American; eGFR = Estimated Glomerular Filtration Rate; Aβ = Amyloid Beta; NfL = Neurofilament Light Chain; GFAP = Glial Fibrillary Acid Protein

Plasma biomarkers were moderately-to-strongly correlated with one another (absolute *rho*=.21-.75; Figure S2) and differed by cognitive diagnosis (CU < MCI < DEM; *p*<.05; Table 1) as previously reported.^1,42^ P-tau181 and p-tau217 were most highly correlated (*rho*=.75; *p*<.001). Plasma biomarkers were also modestly correlated with health-related comorbidities. BMI and eGFR were most strongly negatively correlated with baseline NfL (*rho*: BMI=-.21; eGFR=.39; all *p*<.05) and GFAP (*rho*: BMI=-.26; eGFR=.34; all *p*<.05) levels. Cognition was weakly associated with health-related comorbidities. BMI was positively correlated with memory (*rho*=.15; *p*<.001) and language (*rho*: BMI=.08; *p*<.001) domain z-scores. eGFR was positively correlated with PACC performance (*rho*=.210; *p*<.001). No other associations were observed.

### 3.1 Primary Baseline Cognitive Analyses

#### 3.1.1 General linear models assessing associations between plasma biomarker levels and cognition at baseline

##### 3.1.1.1 Individual baseline models

Figure 1 summarizes relationships between plasma biomarkers and cognitive assessments in partially (Model 1) and fully adjusted (Model 2) models (also see Figure S3). In Model 1, adjusted for baseline age, sex, race, ethnicity, education, and *APOE*-e4 status, baseline plasma p-tau181, p-tau217, NfL, and GFAP were negatively associated with PACC5 performance, and with all cognitive domain z-scores (memory, executive, attention, language, visuospatial) except the phonemic domain (Figure 2, panel a; Table 2). Aβ42/40 levels were positively associated with PACC5 performance and with memory, executive, and language domain z-scores, but not with attention, visuospatial, or phonemic domain z-scores (all *p*>.05; Table 2). In Model 2, additionally adjusted for cardiometabolic factors (baseline BMI, eGFR, hypertension, and diabetes), the association of Aβ42/40 with language was attenuated. In contrast, NfL was newly associated with the phonemic domain (Table 2; Figure 2, panel a). All other associations were unchanged after covariate adjustment and prior to multiple comparisons correction. After correction for multiple comparisons, the p-tau181-phonemic and GFAP-visuospatial associations were no longer statistically significant (Table 2). Independent regression models explained approximately 1-35% of variance in cognitive outcomes assessed (Table S2); the greatest proportion of variance was explained in PACC5 scores.

**Table 2.**
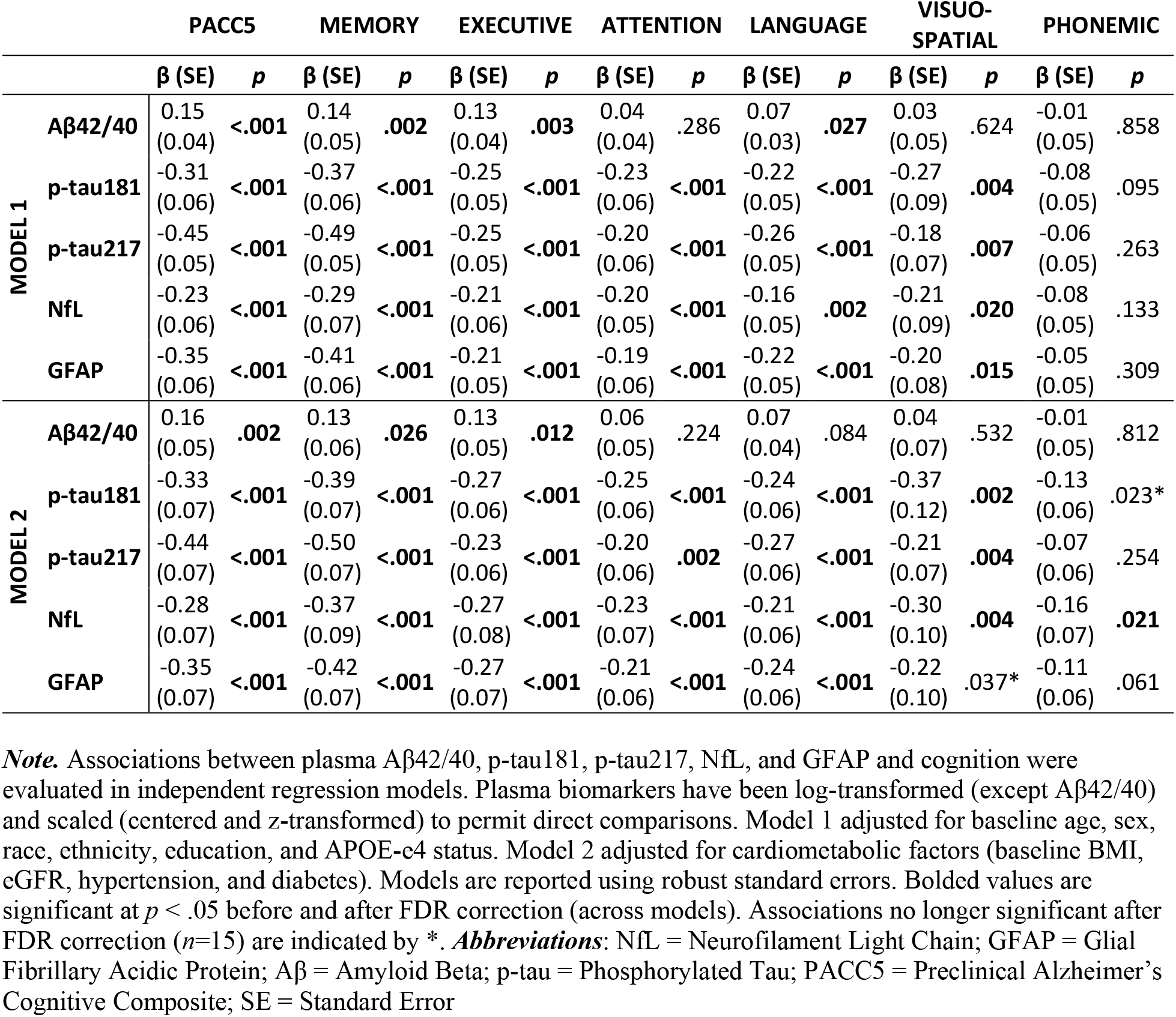
Associations between baseline plasma biomarker levels and cognition at baseline.

|  |  | PACC5 |  | MEMORY |  | EXECUTIVE |  | ATTENTION |  | LANGUAGE |  | VISUO-SPATIAL |  | PHONEMIC |  |
| --- | --- | --- | --- | --- | --- | --- | --- | --- | --- | --- | --- | --- | --- | --- | --- |
| | | $\beta$ (SE) | $p$ | $\beta$ (SE) | $p$ | $\beta$ (SE) | $p$ | $\beta$ (SE) | $p$ | $\beta$ (SE) | $p$ | $\beta$ (SE) | $p$ | $\beta$ (SE) | $p$ |
| MODEL 1 | Aβ42/40 | 0.15<br>(0.04) | **<.001** | 0.14<br>(0.05) | **.002** | 0.13<br>(0.04) | **.003** | 0.04<br>(0.04) | .286 | 0.07<br>(0.03) | **.027** | 0.03<br>(0.05) | .624 | -0.01<br>(0.05) | .858 |
|  | p-tau181 | -0.31<br>(0.06) | **<.001** | -0.37<br>(0.06) | **<.001** | -0.25<br>(0.05) | **<.001** | -0.23<br>(0.06) | **<.001** | -0.22<br>(0.05) | **<.001** | -0.27<br>(0.09) | **.004** | -0.08<br>(0.05) | .095 |
|  | p-tau217 | -0.45<br>(0.05) | **<.001** | -0.49<br>(0.05) | **<.001** | -0.25<br>(0.05) | **<.001** | -0.20<br>(0.06) | **<.001** | -0.26<br>(0.05) | **<.001** | -0.18<br>(0.07) | **.007** | -0.06<br>(0.05) | .263 |
|  | NfL | -0.23<br>(0.06) | **<.001** | -0.29<br>(0.07) | **<.001** | -0.21<br>(0.06) | **<.001** | -0.20<br>(0.05) | **<.001** | -0.16<br>(0.05) | **.002** | -0.21<br>(0.09) | **.020** | -0.08<br>(0.05) | .133 |
|  | GFAP | -0.35<br>(0.06) | **<.001** | -0.41<br>(0.06) | **<.001** | -0.21<br>(0.05) | **<.001** | -0.19<br>(0.06) | **<.001** | -0.22<br>(0.05) | **<.001** | -0.20<br>(0.08) | **.015** | -0.05<br>(0.05) | .309 |
| MODEL 2 | Aβ42/40 | 0.16<br>(0.05) | **.002** | 0.13<br>(0.06) | **.026** | 0.13<br>(0.05) | **.012** | 0.06<br>(0.05) | .224 | 0.07<br>(0.04) | .084 | 0.04<br>(0.07) | .532 | -0.01<br>(0.05) | .812 |
|  | p-tau181 | -0.33<br>(0.07) | **<.001** | -0.39<br>(0.07) | **<.001** | -0.27<br>(0.06) | **<.001** | -0.25<br>(0.06) | **<.001** | -0.24<br>(0.06) | **<.001** | -0.37<br>(0.12) | **.002** | -0.13<br>(0.06) | .023* |
|  | p-tau217 | -0.44<br>(0.07) | **<.001** | -0.50<br>(0.07) | **<.001** | -0.23<br>(0.06) | **<.001** | -0.20<br>(0.06) | **.002** | -0.27<br>(0.06) | **<.001** | -0.21<br>(0.07) | **.004** | -0.07<br>(0.06) | .254 |
|  | NfL | -0.28<br>(0.07) | **<.001** | -0.37<br>(0.09) | **<.001** | -0.27<br>(0.08) | **<.001** | -0.23<br>(0.07) | **<.001** | -0.21<br>(0.06) | **<.001** | -0.30<br>(0.10) | **.004** | -0.16<br>(0.07) | **.021** |
|  | GFAP | -0.35<br>(0.07) | **<.001** | -0.42<br>(0.07) | **<.001** | -0.27<br>(0.07) | **<.001** | -0.21<br>(0.06) | **<.001** | -0.24<br>(0.06) | **<.001** | -0.22<br>(0.10) | .037* | -0.11<br>(0.06) | .061 |

**Note.** Associations between plasma Aβ42/40, p-tau181, p-tau217, NfL, and GFAP and cognition were evaluated in independent regression models. Plasma biomarkers have been log-transformed (except Aβ42/40) and scaled (centered and z-transformed) to permit direct comparisons. Model 1 adjusted for baseline age, sex, race, ethnicity, education, and APOE-ε4 status. Model 2 adjusted for cardiometabolic factors (baseline BMI, eGFR, hypertension, and diabetes). Models are reported using robust standard errors. Bolded values are significant at $p < .05$ before and after FDR correction (across models). Associations no longer significant after FDR correction ( $n=15$ ) are indicated by \*. **Abbreviations:** NfL = Neurofilament Light Chain; GFAP = Glial Fibrillary Acidic Protein; Aβ = Amyloid Beta; p-tau = Phosphorylated Tau; PACC5 = Preclinical Alzheimer's Cognitive Composite; SE = Standard Error

**Figure 1.**
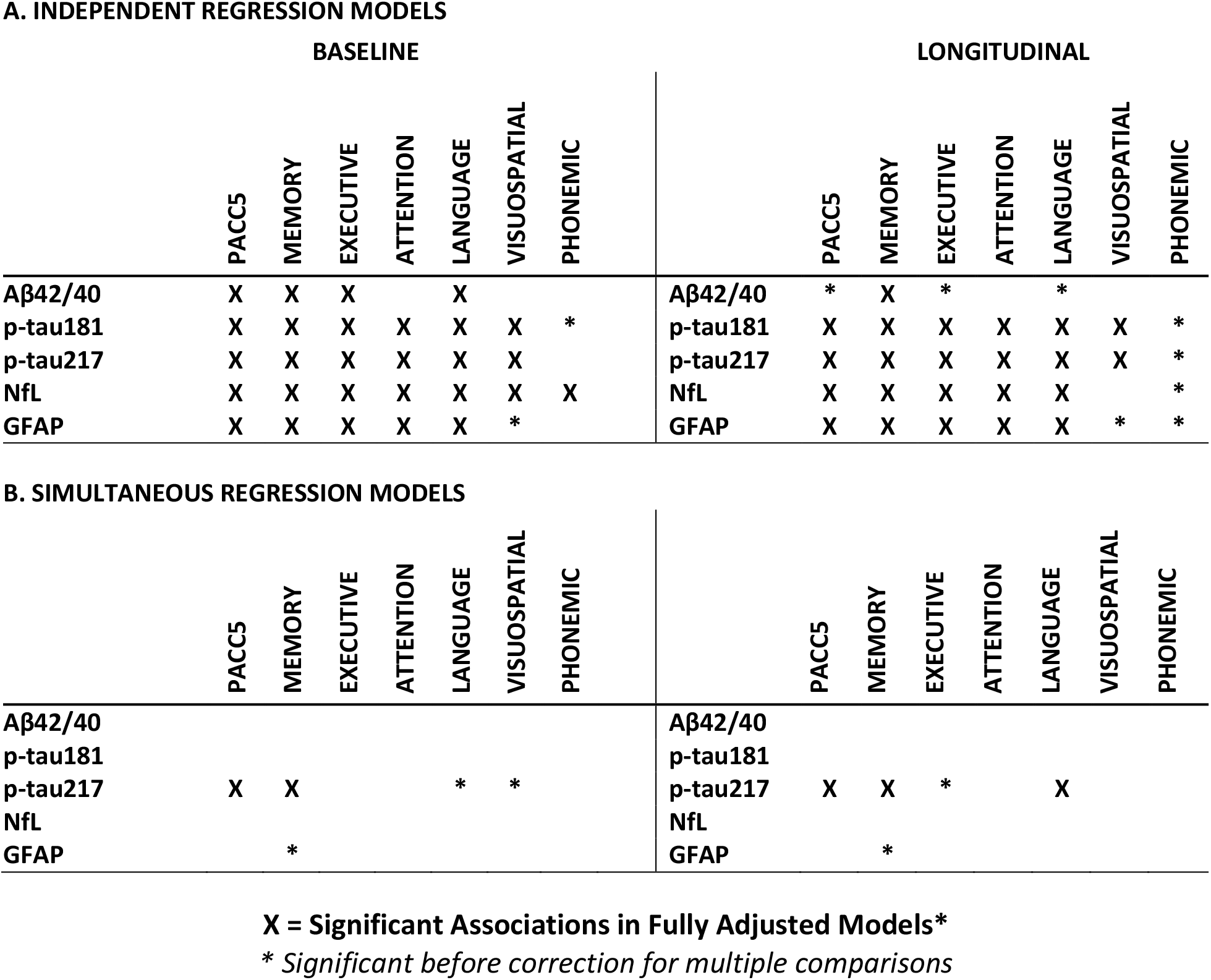
Graphical summary highlighting robust associations between altered baseline plasma biomarker levels and poorer cognitive performance. ***Note:*** Graphical summary of robust associations between baseline plasma biomarkers and cognitive performance. Statistically significant associations (X) are visually depicted for fully adjusted (age, sex, race, ethnicity, education, APOE-ε4 status, estimated glomerular filtration rate, body mass index, diabetes, and hypertension) independent and simultaneous (e.g., all baseline plasma biomarkers entered together) regression models after correction for multiple comparisons. This figure provides a qualitative overview of where associations are observed across modeling approaches and does not convey effect size magnitude, relative strength, or the degree of attenuation versus stability between independent and simultaneous models; these details are reported in the corresponding tables. ***Abbreviations***: NfL = Neurofilament Light Chain; GFAP = Glial Fibrillary Acidic Protein; Aβ = Amyloid Beta; p-tau = Phosphorylated Tau; PACC5 = Preclinical Alzheimer’s Cognitive Composite

**Figure 2.**
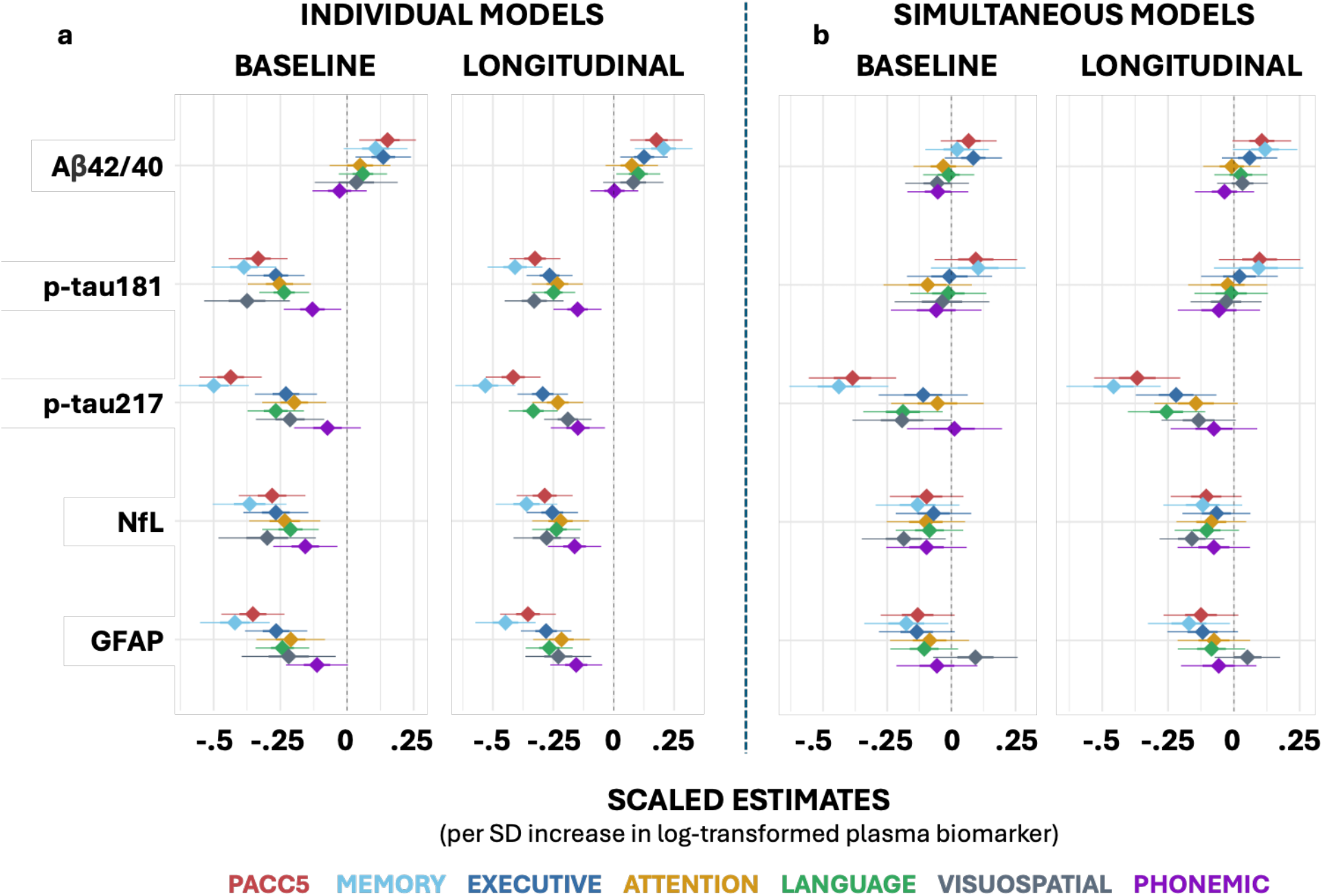
Associations between baseline plasma biomarker levels and cognitive outcomes at baseline and over time in independent and simultaneous regression models. ***Note***. Associations between plasma Aβ42/40, p-tau181, p-tau217, NfL, and GFAP and cognition were evaluated using independent (left panels) and simultaneous (e.g., all plasma biomarkers entered together; right panels) multivariable regression models adjusted for demographic and health factors. Here, coefficients from fully adjusted models (Model 2) are displayed. These models were adjusted for baseline age, sex, race, ethnicity, education, eGFR, BMI, diabetes, and hypertension. Models evaluating longitudinal change in cognition were additionally adjusted for time between visits. Scaled estimates are shown, as plasma biomarkers were log-transformed (except Aβ42/40) and standardized (centered and z-transformed) prior to analysis to aid visualization and permit direct comparison across biomarkers. ***Abbreviations***: NfL = Neurofilament Light Chain; GFAP = Glial Fibrillary Acidic Protein; Aβ = Amyloid Beta; p-tau = Phosphorylated Tau; PACC5 = Preclinical Alzheimer’s Cognitive Composite.

##### 3.1.1.2 Simultaneous baseline models

Additional analyses were conducted to examine the associations of plasma biomarkers (Aβ42/40, p-tau181, p-tau217, NfL, and GFAP) and cognition when all biomarkers were entered into the same models simultaneously (Models 1 and 2; see Methods; Figure 1). Across simultaneous baseline analyses, p-tau217 was negatively associated with PACC5 performance and with memory, executive, language, and visuospatial domain z-scores in partially adjusted models (Model 1); however, after full adjustment (Model 2) and following correction for multiple comparisons, robust associations persisted only between p-tau217 and PACC5 or memory (Figure 2, panel a; Table S3). GFAP showed nominal associations with memory that did not survive correction for multiple comparisons (Figure 1; Table S3). Notably, when all biomarkers were entered simultaneously, p-tau217 was the only biomarker that remained significantly associated with cognitive outcomes (Figure 2, panel a; Table S3). Simultaneous regression models explained approximately 3-37% of variance in cognitive outcomes assessed (Table S3), with plasma biomarkers explaining the greatest variance in PACC5 performance.

### 3.2 Primary Longitudinal Analyses

#### 3.2.1 Longitudinal mixed effect models assessing associations between baseline plasma biomarker levels and average change in cognitive performance

##### 3.2.1.1 Independent longitudinal models

Figure 1 summarizes significant relationships between plasma biomarkers and cognition across models. Figure 2 summarizes associations between plasma biomarker levels and change in cognition over time in partially (Model 1) and fully adjusted (Model 2) models (also see Figure S4). In Model 1, lower baseline Aβ42/40 and higher p-tau181, p-tau217, NfL, and GFAP levels were associated with declines in cognitive performance on the PACC5 and memory, executive, and language domain z-scores (*all p*<.05; Figure 2, panel b; Table 3). All plasma biomarkers except for Aβ42/40 were associated with greater decline in attention and visuospatial domain z-scores (Table 3). All other associations were preserved after covariate adjustment (i.e. Model 2). Only p-tau217 was associated with greater decline in phonemic domain z-scores after FDR correction (Table 3). Independent regression models explained approximately 1-34% of [marginal] variance in cognitive outcome, with p-tau217 accounting for the greatest variance in PACC5 scores (Table S4).

**Table 3.** Longitudinal associations between baseline plasma biomarker levels and cognition at follow-up.

|  |  | PACC5 |  | MEMORY |  | EXECUTIVE |  | ATTENTION |  | LANGUAGE |  | VISUO-SPATIAL |  | PHONEMIC |  |
| --- | --- | --- | --- | --- | --- | --- | --- | --- | --- | --- | --- | --- | --- | --- | --- |
| | | $\beta$ (SE) | *p* | $\beta$ (SE) | *p* | $\beta$ (SE) | *p* | $\beta$ (SE) | *p* | $\beta$ (SE) | *p* | $\beta$ (SE) | *p* | $\beta$ (SE) | *p* |
| **MODEL 1** | **Aβ42/40** | 0.15<br>(0.04) | **<.001** | 0.20<br>(0.05) | **<.001** | 0.11<br>(0.04) | .009* | 0.05<br>(0.05) | 0.27 | 0.11<br>(0.04) | .005* | 0.06<br>(0.05) | 0.24 | 0.01<br>(0.04) | 0.74 |
|  | **p-tau181** | -0.30<br>(0.04) | **<.001** | -0.39<br>(0.05) | **<.001** | -0.25<br>(0.04) | **<.001** | -0.25<br>(0.04) | **<.001** | -0.21<br>(0.04) | **<.001** | -0.27<br>(0.05) | **<.001** | -0.09<br>(0.04) | .029* |
|  | **p-tau217** | -0.42<br>(0.04) | **<.001** | -0.54<br>(0.05) | **<.001** | -0.30<br>(0.04) | **<.001** | -0.26<br>(0.04) | **<.001** | -0.31<br>(0.04) | **<.001** | -0.18<br>(0.04) | **<.001** | -0.12<br>(0.04) | .009* |
|  | **NfL** | -0.23<br>(0.05) | **<.001** | -0.30<br>(0.05) | **<.001** | -0.20<br>(0.04) | **<.001** | -0.21<br>(0.05) | **<.001** | -0.18<br>(0.04) | **<.001** | -0.19<br>(0.06) | .001* | -0.08<br>(0.05) | 0.07 |
|  | **GFAP** | -0.34<br>(0.05) | **<.001** | -0.44<br>(0.05) | **<.001** | -0.23<br>(0.04) | **<.001** | -0.22<br>(0.05) | **<.001** | -0.23<br>(0.04) | **<.001** | -0.23<br>(0.06) | **<.001** | -0.07<br>(0.05) | 0.11 |
| **MODEL 2** | **Aβ42/40** | 0.18<br>(0.06) | .001* | 0.22<br>(0.06) | **<.001** | 0.12<br>(0.05) | .015* | 0.09<br>(0.06) | 0.1 | 0.11<br>(0.05) | .018* | 0.10<br>(0.06) | 0.13 | 0.01<br>(0.05) | 0.83 |
|  | **p-tau181** | -0.33<br>(0.05) | **<.001** | -0.41<br>(0.06) | **<.001** | -0.27<br>(0.05) | **<.001** | -0.23<br>(0.05) | **<.001** | -0.25<br>(0.05) | **<.001** | -0.33<br>(0.06) | **<.001** | -0.15<br>(0.05) | .003* |
|  | **p-tau217** | -0.42<br>(0.06) | **<.001** | -0.53<br>(0.06) | **<.001** | -0.29<br>(0.05) | **<.001** | -0.23<br>(0.05) | **<.001** | -0.33<br>(0.05) | **<.001** | -0.19<br>(0.05) | **<.001** | -0.15<br>(0.06) | .010* |
|  | **NfL** | -0.29<br>(0.06) | **<.001** | -0.36<br>(0.06) | **<.001** | -0.26<br>(0.05) | **<.001** | -0.22<br>(0.06) | **<.001** | -0.24<br>(0.05) | **<.001** | -0.28<br>(0.07) | **<.001** | -0.16<br>(0.06) | .004* |
|  | **GFAP** | -0.36<br>(0.06) | **<.001** | -0.45<br>(0.06) | **<.001** | -0.28<br>(0.05) | **<.001** | -0.22<br>(0.06) | **<.001** | -0.27<br>(0.05) | **<.001** | -0.23<br>(0.07) | .001* | -0.16<br>(0.05) | .005* |

**Note.** Longitudinal mixed models examined associations between baseline plasma levels of Aβ42/40, p-tau181, p-tau217, NfL, and GFAP and changes in cognition overtime. Plasma biomarkers (except Aβ42/40) were log- and scaled (centered and z-transformed) to permit direct comparisons. Model 1 adjusted for baseline age, sex, race, ethnicity, education, and APOE-e4 status. Model 2 adjusted for cardiometabolic factors (baseline BMI, eGFR, hypertension, and diabetes). Models are reported using robust standard errors. Bolded values are significant at $p < .05$ before and after FDR correction (across models). Associations no longer significant after FDR correction ( $n=15$ ) are indicated by \*. **Abbreviations:** NfL = Neurofilament Light Chain; GFAP = Glial Fibrillary Acidic Protein; Aβ = Amyloid Beta; p-tau = Phosphorylated Tau; PACC5 = Preclinical Alzheimer's Cognitive Composite; SE = Standard Error

##### 3.2.1.2 Simultaneous longitudinal models

Additional analyses included all baseline plasma biomarkers in the same models. In Model 1, baseline Aβ42/40 was associated with memory domain z-scores; however, this association was attenuated in Model 2 (Table S5). As with the baseline simultaneous models, p-tau217 was most consistently associated with cognitive decline when all biomarkers were entered simultaneously. Higher baseline p-tau217 was associated with declines in PACC5 performance and all cognitive domain z-scores (all nominal *p*<.05) except for the visuospatial and phonemic domains (Figure 2, panel b; Table S5). Higher baseline GFAP was associated with greater decline in PACC5 in partially adjusted models (Model 1), but this association was attenuated and no longer statistically significant after additional covariate adjustment in Model 2 (Figure 2a; Table S5). Higher GFAP was also associated with decline in memory domain z-scores in Model 1 and remained significantly associated, though attenuated, in Model 2; however, these associations did not survive correction for multiple comparisons (Figure 2b; Table S5).

### 3.3 Post-hoc analyses

Overall, in fully adjusted models, evidence of effect modification by baseline cognitive status (CU vs MCI; see Methods) on plasma biomarker–cognition associations was sparse or domain-specific. In cross-sectional analyses, assessed independently, cognitive status modified associations between p-tau181 or NfL and phonemic domain z-scores. When assessed simultaneously, modification was observed only for the association between NfL and executive domain z-scores. Longitudinally, baseline cognitive status modified associations between p-tau181, p-tau217, or GFAP and PACC5 and memory domain z-scores, and for A?42/40 and language domain z-scores (all p < 0.05; Table S6). In these models, higher biomarker burden was generally associated with poorer cognitive performance, with steeper negative biomarker–cognition associations observed among participants with MCI compared with CU, indicating more pronounced associations in the MCI group. In simultaneous longitudinal mixed-effects models accounting for shared biological variance across plasma biomarkers, interaction effects comparing CU vs MCI were attenuated across domains with the exception of the association between GFAP and memory domain z-score (Table S6).

## 4 Discussion

Plasma biomarkers have emerged as minimally invasive and cost-effective tools for detecting AD/ADRD pathology^1–5^ and have been increasingly associated with concurrent and emerging cognitive impairment.^12,15–31^ Building on this foundational work, we sought to clarify the relationship between the plasma ADRD biomarkers with global and domain-specific cognition in a community-dwelling cohort with a high prevalence of cardiometabolic comorbidities, a population that remains underrepresented in biomarker studies despite its clinical relevance. Accordingly, we examined whether baseline plasma ADRD biomarker levels were associated with global and domain-specific cognitive performance both cross-sectionally and longitudinally (up to 8 years; mean follow-up ∼4 years). Across this heterogeneous cohort, higher levels of p-tau181, p-tau217, NfL, and GFAP and a lower Aβ42/40 ratio were associated with poorer cognitive performance on the PACC5 preclinical composite and across all cognitive domains except phonemic fluency. These associations were overall robust to adjustment for demographic factors, APOE ε4 status, comorbidities, cardiometabolic health factors, and cognitive status, and were observed in both cross-sectional and longitudinal mixed-effects models.

A central aim of this study was to characterize how plasma biomarkers relate to both preclinical cognitive composites and domain-specific cognitive performance. We found that a lower Aβ42/40 ratio was associated with poorer PACC5 performance and with lower memory, executive function, and language z-scores. These findings align with prior work demonstrating associations between Aβ42/40 ratio and cognitive performance, particularly in memory-related domains.^16,30^ However, variability across studies may reflect differences in cohort composition, comorbidity burden, cognitive measures, or assay platforms. We also observed that higher levels of p-tau181, p-tau217, NfL, and GFAP were associated with poorer PACC5 performance and with lower memory, executive, language, attention, and visuospatial domain z-scores. Among these biomarkers, p-tau217 consistently showed the strongest associations with PACC5 and memory performance, supporting its sensitivity to early memory changes captured by composite measures, consistent with prior work.^25,67^ Notably, plasma biomarkers collectively explained more variance in PACC5 performance than in any single cognitive domain, even after covariate adjustment, highlighting the sensitivity of composite outcomes to ADRD plasma biomarker-related cognitive change.

Although prior studies have shown that p-tau217 outperforms other plasma biomarkers in predicting incident AD dementia,^68^ fewer investigations have examined how plasma biomarkers perform when evaluated simultaneously in relation to preclinical cognitive composites and domain-specific outcomes. To address this gap, we evaluated models in which all baseline plasma biomarkers were entered simultaneously. Across these models, p-tau217 emerged as the most informative independent predictor of baseline cognitive performance. Specifically, p-tau217 was associated with PACC5 performance and with memory, executive, language, and visuospatial domain z-scores in partially adjusted models (Model 1). After further adjustment for cardiometabolic factors (BMI, eGFR, hypertension, diabetes; Model 2) and correction for multiple comparisons, robust associations for p-tau217 persisted for global cognition (PACC5) and memory. This pattern suggests that cardiometabolic health may differentially influence biomarker–cognition relationships across cognitive domains, potentially contributing to heterogeneity across studies.

Longitudinally, these same baseline plasma biomarkers were associated with change in PACC5 performance and in memory, executive, and language domains, and, except for Aβ42/40, with change in attention and visuospatial performance. In simultaneous longitudinal models, elevated p-tau217 again emerged as the strongest and most consistent predictor of cognitive decline, whereas associations for other biomarkers were attenuated. When all baseline plasma biomarkers were modeled simultaneously, p-tau217 most consistently retained an independent association with longitudinal cognitive change. Importantly, independent and simultaneous models address complementary questions: independent models capture overall biomarker–cognition associations, whereas simultaneous models assess whether these associations persist when biomarkers are considered simultaneously. Attenuation in simultaneous models therefore reflects shared variance within a conservative inferential framework and potentially convergent biological pathways, rather than absence of biological relevance. These findings reinforce the central role of p-tau217 as a sensitive indicator of emerging decline, while also suggesting that broader biomarker panels may be informative for identifying individuals with multidomain or atypical cognitive impairment, often observed in non-AD dementias^48,49,70^ or for improving prediction of progression from MCI to AD dementia over four years.^69^

Post-hoc analyses further examined whether cognitive status modified biomarker-cognition associations. Associations of p-tau181 and p-tau217 with PACC5 performance were more pronounced among participants with MCI compared with cognitively unimpaired individuals. Although the PACC5 was developed to detect subtle amyloid-related decline in preclinical AD, stronger biomarker-cognition associations in MCI are expected, as individuals in this transitional stage exhibit greater cognitive variability and higher underlying pathological burden. These findings are consistent with prior work demonstrating that cognitive composites are sensitive to early decline while also capturing more pronounced associations as individuals progress from preclinical to prodromal stages of AD-dementia.^48,49,70^

### 4.1 Strengths & Limitations

A key strength of this study is the inclusion of a large, clinically heterogeneous cohort with a high prevalence of cardiometabolic comorbidities, enhancing the real-world relevance of findings as plasma biomarkers move toward broader clinical implementation. Approximately 60% of older adults with cognitive impairment have three or more chronic conditions,^71^ underscoring the importance of evaluating plasma biomarker performance in such populations. There are also potential limitations that warrant discussion. First, the predominance of CU and MCI participants aligns with the WFADRC mission to study early cognitive change. However, this focus may limit generalizability to individuals with more advanced cognitive impairment. Second, high between-participant variability, modest within-participant change, and irregular follow-up constrained the added value of more complex random-effects structures. Accordingly, models incorporated participant-level random intercepts, which adequately captured the longitudinal correlation structure specific to the study’s primary aims. Finally, baseline plasma biomarkers explained a modest proportion of variance in cognitive outcomes, and substantial unexplained variability remained even in fully adjusted models. The plasma biomarkers evaluated here represent a portion of the pathology underlying ADRD. Future work integrating longitudinal biomarker trajectories, multimodal data, additional ADRD and other biomarkers, and cross-cohort replication will be critical for refining risk stratification and understanding heterogeneity in cognitive aging, given the limited number of cases progressing to either MCI or dementia within a single cohort. Although the present study was limited to baseline plasma biomarker measures, longitudinal plasma biomarker collection is underway in the WFADRC and will enable future evaluation of biomarker dynamics in relation to clinical progression.

### 4.2 Conclusion

In a heterogeneous, community-dwelling cohort, pathologic plasma biomarker levels, particularly p-tau217, were associated with poorer baseline cognition and greater decline across multiple domains, largely independent of demographic and clinical factors. Simultaneous modeling demonstrated that p-tau217 contributed the most conditionally independent information about cognitive impairment, with other biomarkers reflecting related AD/ADRD-linked processes. These findings illustrate the utility of multimarker approaches for parsing shared versus biomarker-specific associations in real-world cohorts. Future longitudinal analyses will evaluate whether within-person biomarker changes and evolving comorbidities are associated with clinical progression.

## Acknowledgements

This work was supported by the Wake Forest University School of Medicine’s Alzheimer’s Disease Research Center (P30AG049638, P30AG072947, R01AG054069, R01AG058969, and T32AG033534), which is funded by the National Institute on Aging (NIA). Additional support was provided by the Department of Gerontology and Geriatric Medicine and Center for Healthy Aging and Alzheimer’s Prevention, Wake Forest School of Medicine. Plasma biomarker analyses were completed in part by the NCRAD Biomarker Assay Laboratory as part of the Alzheimer’s Disease Center Fluid Biomarker (ADCFB) Initiative, which receives government support under a cooperative agreement grant (U24 AG021886) awarded by the National Institute on Aging (NIA). Plasma p-tau217 analytes were processed at Neurocode (Bellingham, WA) and funded by ALZpath (Carlsbad CA). This study would not have been possible without the commitment and support of our valued WFADRC staff and study participants.

## Consent Statement

Written informed consent was obtained for all participants and/or their legally authorized representatives.

## Data Availability

Data are available upon reasonable request and can be obtained via WakeShare.org.

