## Supplemental Material for "Plasma biomarker levels and cognitive decline in a heterogenous community-based cohort with multiple comorbidities"

**Supplemental Materials**

**Figure S1. Summary of available cognitive assessments across clinical visits in the WF ADRC**

**Summary overview of datapoints by year and baseline cognitive status**

| | TOTAL | 1 | 2 | 3 | 4 | 5 | 6 | 7 | 8 |
|---|---|---|---|---|---|---|---|---|---|
| TOTAL | 2,389 | 590 | 444 | 345 | 307 | 269 | 247 | 116 | 71 |
| CU | 1,463 (61.2%) | 314 (53.2%) | 261 (58.8%) | 208 (60.3%) | 206 (67.1%) | 170 (63.2%) | 165 (66.8%) | 82 (70.7%) | 57 (80.3%) |
| MCI | 758 (31.7%) | 206 (34.9%) | 144 (32.4%) | 112 (32.5%) | 89 (29.0%) | 88 (32.7%) | 75 (30.4%) | 31 (26.7%) | 13 (18.3%) |
| DEM | 168 (7.0%) | 70 (11.9%) | 39 (8.8%) | 25 (7.2%) | 12 (3.9%) | 11 (4.1%) | 7 (2.8%) | 3 (2.6%) | 1 (1.4%) |

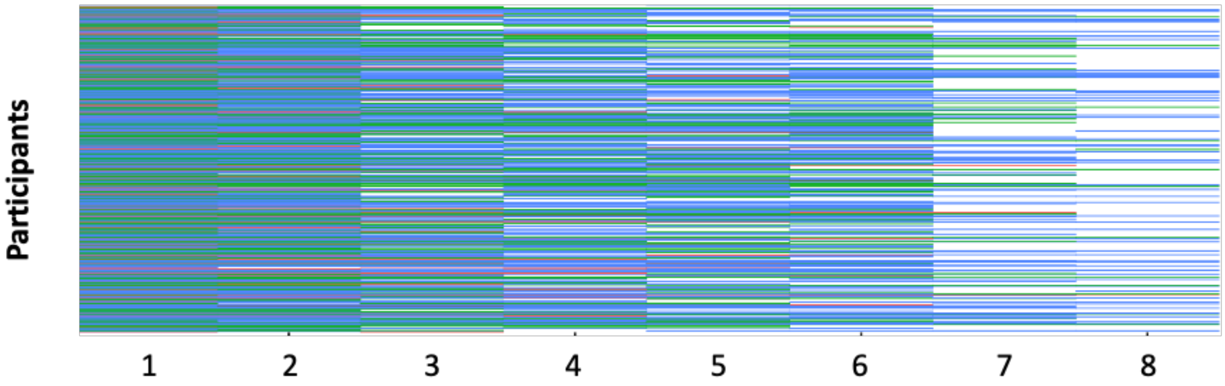


***Note.*** Participants in the WF ADRC with baseline plasma biomarker data had up to 7 years of follow-up cognitive assessments (Session 1 = baseline). Approximately 70% of the cohort completed three or more visits, with an average interval of 1,221 days (3.3 years) between the first and last cognitive assessment. ***Abbreviations*:** CU = Cognitively Unimpaired; DEM = Dementia; MCI = Mild Cognitive Impairment; WF ADRC = Wake Forest Alzheimer's Disease Research Center.

**Figure S2. Unadjusted spearman correlations between baseline plasma biomarker levels and baseline cognition**

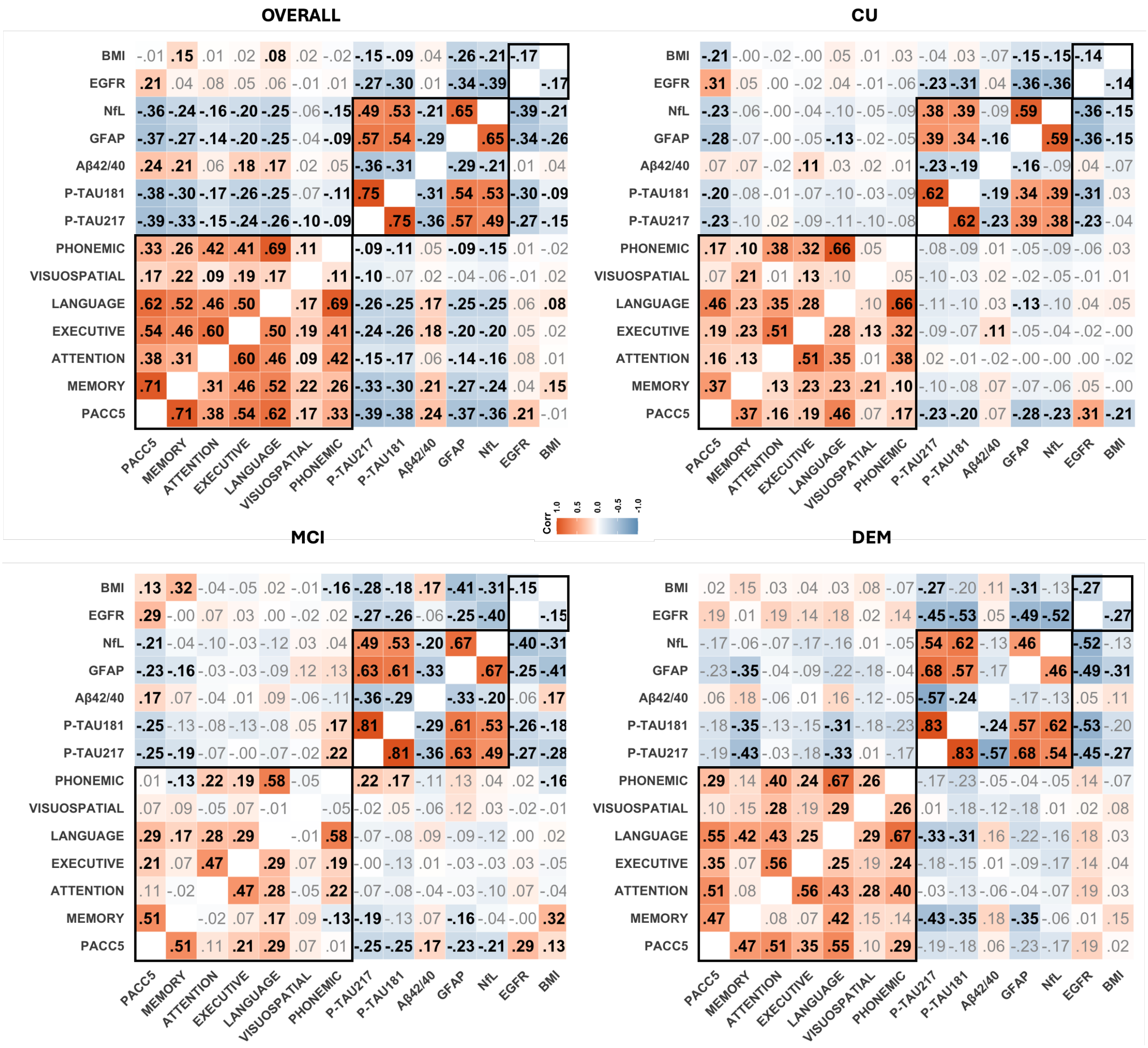


***Note*****:** Unadjusted Spearman rank correlations assessing relationships between baseline plasma biomarkers and cognition. Plasma biomarkers and cognitive outcomes were moderately to highly correlated (absolute *rho*: .29-.57). Within each modality, p-tau181 and p-tau217 (*rho* = .75) and PACC5 performance and memory domain scores (*rho* = .71) were most strongly correlated. P-tau217 showed the strongest correlation with poorer PACC5 performance and memory domain z-scores (*rho* = -.39 and -.33). Significant values are displayed in bold black text. ***Abbreviations***: Aβ = Amyloid Beta; CU = Cognitively Unimpaired; DEM.= Dementia; GFAP = Glial Fibrillary Acidic Protein; MCI = Mild Cognitive Impairment; NfL = Neurofilament Light Chain; PACC5 = Preclinical Alzheimer's Cognitive Composite; p-tau = Phosphorylated Tau.

**Figure S3. Unadjusted relationships between baseline plasma biomarkers and cognitive performance at baseline**

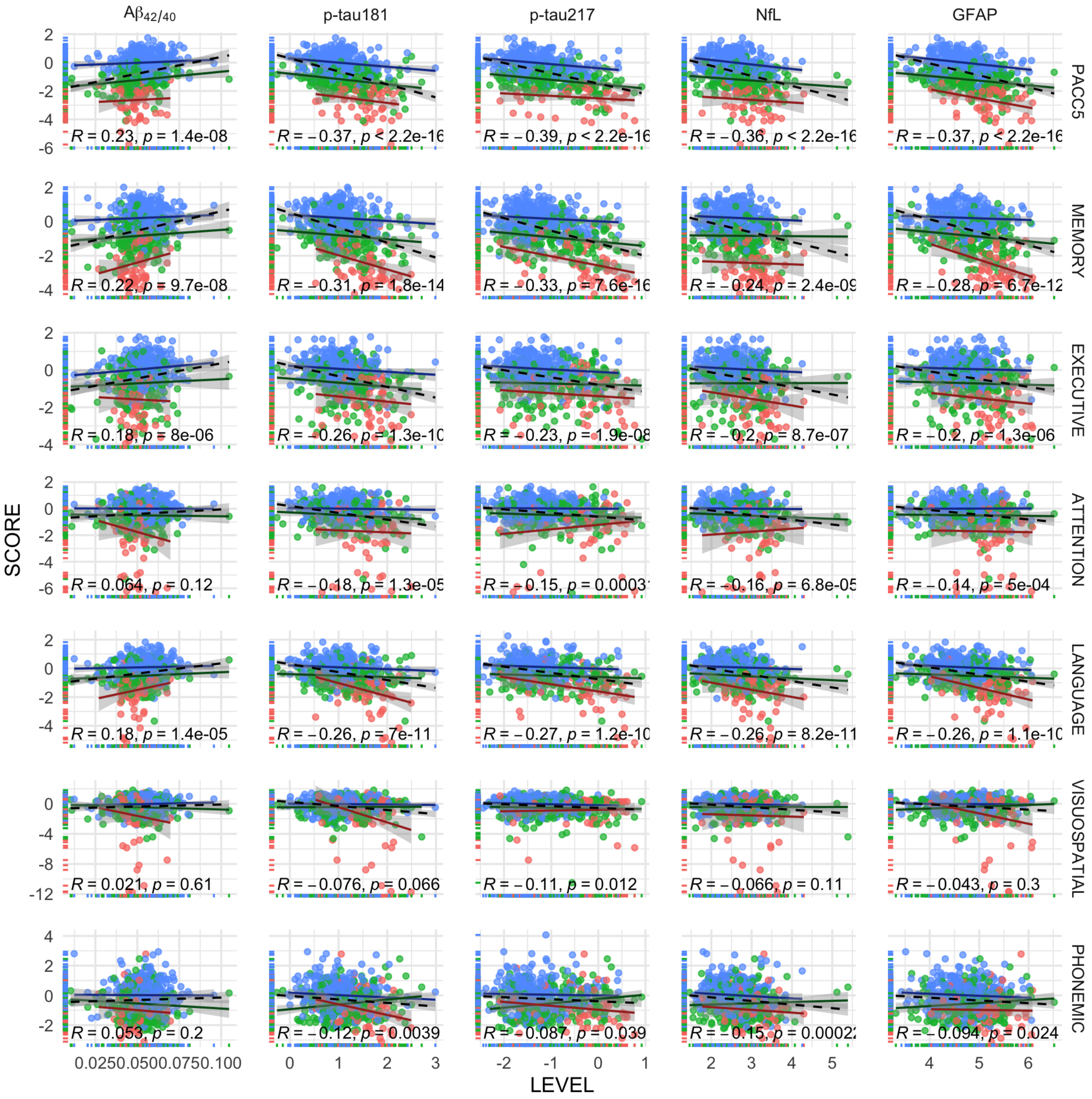


***Note***. Scatterplots depict unadjusted associations between baseline plasma biomarker levels and baseline cognitive performance across domains in the full sample (black) and stratified by baseline cognitive status: cognitively unimpaired (CU; blue), mild cognitive impairment (MCI; green), and dementia (DEM; red). Each point represents an individual participant. Solid lines indicate group-specific linear fits, with Spearman correlation coefficients and corresponding *p*-values shown for descriptive purposes for the full sample. ***Abbreviations***: Aβ = Amyloid Beta; CU = Cognitively Unimpaired; DEM = Dementia; GFAP = Glial Fibrillary Acidic Protein; MCI = Mild Cognitive Impairment; NfL = Neurofilament Light Chain; PACC5 = Preclinical Alzheimer's Cognitive Composite; p-tau = Phosphorylated Tau.

**Figure S4. Correlation of PACC5 performance between baseline and follow-up visits**

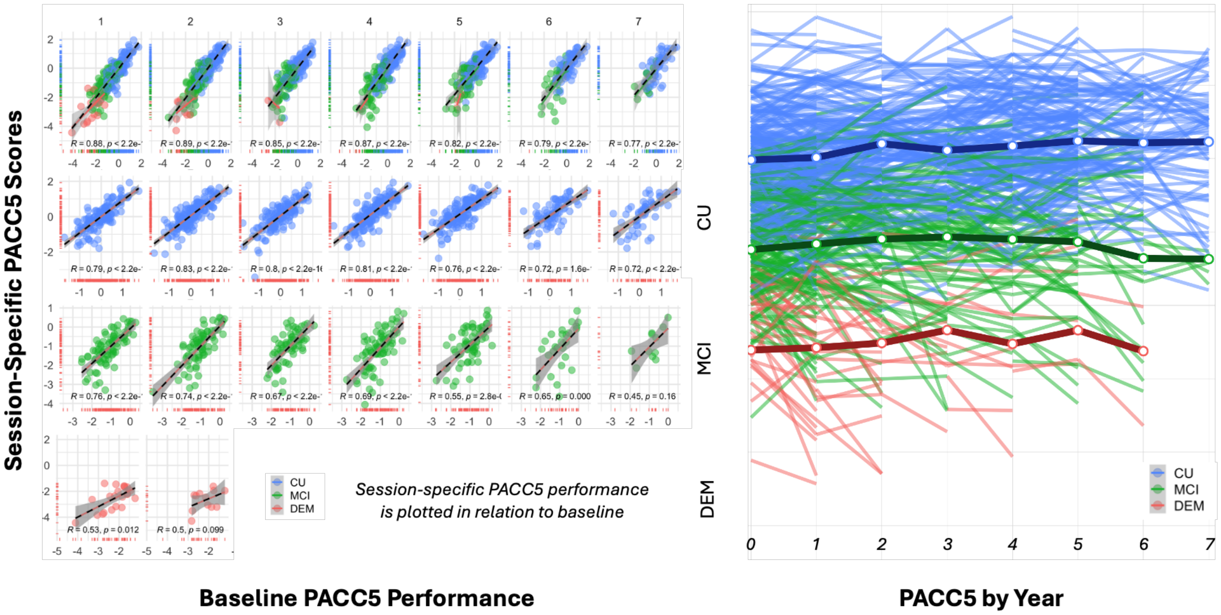


***Note.*** *Left panel:* Scatterplots depict unadjusted relationships between baseline PACC5 performance and session-specific PACC5 scores across follow-up visits in the full analytic sample (top row) and stratified by baseline cognitive status: cognitively unimpaired (CU; second row), mild cognitive impairment (MCI; third row), and dementia (DEM; fourth row). Each point represents an individual participant at a given visit, with participants contributing multiple observations across sessions. These plots illustrate the high within-participant correlation in cognitive performance across visits. *Right panel:* Spaghetti plot showing unadjusted longitudinal trajectories of PACC5 performance for individual participants over time, stratified by baseline cognitive status. ***Abbreviations***: Aβ = Amyloid Beta; CU = Cognitively Unimpaired; DEM = Dementia; GFAP = Glial Fibrillary Acidic Protein; MCI = Mild Cognitive Impairment; NfL = Neurofilament Light Chain; PACC5 = Preclinical Alzheimer's Cognitive Composite; p-tau = Phosphorylated Tau.

**Table S1. Baseline participants characteristics by diagnostic and overall conversion groups**

| | | | | CONVERSION BY DIAGNOSTIC GROUP | | | COMPARISONS BY OVERALL CONVERSION STATUS STABLE VS CONVERTER GROUPS | | | | |
|---|---|---|---|---|---|---|---|---|---|---|---|
| | | N | Overall N = 590[1] | CU-MCI N = 25[1] | CU-DEM N = 3[1] | MCI-DEM N = 32[1] | STABLE N = 530[1] | CONVERTER N = 60[1] | d[2] | 95% CI[2,3] | p-FDR[3] |
| Demographics and Health Factors | Age | 590 | 70 (8) | 74 (9) | 80 (8) | 74 (6) | 70 (8) | 74 (7) | **0.55** | **0.28, 0.82** | **<.001** |
| | Race | 590 | | | | | | | 0.25 | -0.02, 0.51 | .400 |
| | W | | 473 (80%) | 20 (80%) | 3 (100%) | 29 (91%) | 421 (79%) | 52 (87%) | | | |
| | B/AA | | 110 (19%) | 0 (0%) | 0 (0%) | 0 (0%) | 103 (19%) | 7 (12%) | | | |
| | A | | 5 (0.8%) | 1 (4.0%) | 0 (0%) | 0 (0%) | 4 (0.8%) | 1 (1.7%) | | | |
| | AI/AN | | 2 (0.3%) | 4 (16%) | 0 (0%) | 3 (9.4%) | 2 (0.4%) | 0 (0%) | | | |
| | Female | | 382 (65%) | 19 (76%) | 3 (100%) | 18 (56%) | 342 (65%) | 40 (67%) | 0.05 | -0.22, 0.31 | .800 |
| | Education | 590 | 15.77 (2.51) | 15.08 (2.61) | 13.67 (2.89) | 16.09 (2.86) | 15.80 (2.48) | 15.55 (2.79) | -0.1 | -0.37, 0.17 | .500 |
| | APOE4 (Carrier) | 576 | 193 (34%) | 4 (16%) | 1 (33%) | 16 (50%) | 172 (33%) | 21 (35%) | 0.04 | -0.23, 0.30 | .800 |
| | BMI | 590 | 27.9 (5.6) | 29.3 (6.6) | 27.4 (3.9) | 25.7 (4.5) | 27.9 (5.6) | 27.3 (5.7) | -0.1 | -0.38, 0.16 | .400 |
| | eGFR | **411** | **79 (15)** | **72 (14)** | **61 (NA)** | **74 (17)** | **79 (15)** | **73 (16)** | **-0.4** | **-0.73, -0.11** | **.022** |
| | Pre/Diabetes | 590 | 377 (64%) | 18 (72%) | 3 (100%) | 24 (75%) | 332 (63%) | 45 (75%) | 0.28 | 0.02, 0.55 | .200 |
| | HTN | 590 | 257 (44%) | 13 (52%) | 1 (33%) | 15 (47%) | 229 (43%) | 28 (47%) | 0 | -0.27, 0.27 | .700 |
| Plasma Biomarkers | Aβ42/40 | 586 | 0.053 (0.011) | 0.053 (0.007) | 0.045 (0.013) | 0.044 (0.012) | 0.053 (0.010) | 0.048 (0.011) | **-0.6** | **-0.83, -0.29** | **.001** |
| | p-tau181 | 590 | 3.46 (2.00) | 2.98 (1.23) | 4.50 (0.94) | 5.28 (2.41) | 3.37 (1.96) | 4.28 (2.23) | **0.46** | **0.19, 0.73** | **.001** |
| | p-tau217 | 503 | 0.43 (0.36) | 0.38 (0.21) | 0.76 (0.42) | 0.87 (0.54) | 0.41 (0.34) | 0.65 (0.48) | **0.69** | **0.41, 0.97** | **<.001** |
| | NfL | 586 | 17 (14) | 17 (7) | 19 (2) | 21 (7) | 17 (15) | 19 (7) | **0.15** | **-0.12, 0.41** | **<.001** |
| | GFAP | 586 | 137 (76) | 135 (63) | 158 (11) | 197 (86) | 133 (75) | 169 (80) | **0.48** | **0.21, 0.75** | **<.001** |
| Cognition | PACC5 | 574 | -0.71 (1.13) | -0.70 (0.58) | -0.74 (0.42) | -1.64 (0.60) | -0.66 (1.15) | -1.20 (0.75) | **-0.5** | **-0.75, -0.20** | **<.001** |
| | Memory | 590 | -0.47 (1.16) | -0.17 (0.53) | -0.32 (0.63) | -1.33 (0.93) | -0.43 (1.17) | -0.80 (0.96) | **-0.3** | **-0.58, -0.05** | **.010** |
| | Executive | 590 | -0.40 (0.98) | -0.23 (0.59) | -0.21 (0.84) | -1.09 (1.14) | -0.37 (0.97) | -0.69 (1.02) | **-0.3** | **-0.60, -0.06** | **.017** |
| | Attention | 590 | -0.38 (1.09) | -0.16 (0.61) | -0.53 (0.19) | -0.73 (0.82) | -0.37 (1.12) | -0.48 (0.76) | **-0.1** | **-0.38, 0.16** | **.110** |
| | Language | 590 | -0.32 (0.89) | -0.28 (0.58) | -0.21 (0.27) | -0.91 (0.80) | -0.29 (0.90) | -0.61 (0.76) | **-0.4** | **-0.63, -0.10** | **.002** |
| | Visuospatial | 590 | -0.35 (1.42) | -0.36 (0.74) | 0.28 (0.93) | -0.54 (1.10) | -0.34 (1.47) | -0.42 (0.96) | **-0.1** | **-0.32, 0.21** | **.200** |
| | Phonemic | 588 | -0.30 (0.99) | -0.26 (0.78) | -0.05 (0.85) | -0.64 (0.79) | -0.28 (1.01) | -0.45 (0.80) | **-0.2** | **-0.44, 0.09** | **.400** |

***Note***: Approximately 10% (n=60) of participants converted to MCI or DEM during the course of the study. Compared to those who remained stable in their baseline diagnosis, converters were (1) older and had lower eGFR on average; (2) had more pathologic plasma biomarker levels, p-tau217 in particular; and (3) had poorer cognition across assessments with PACC5 best distinguishing the groups. ***Abbreviations***: AA = African American; Aβ = Amyloid Beta; AI/AN = American Indian or Alaskan Native; CU = Cognitively Normal; DEM = Dementia; eGFR = Estimated Glomerular Filtration Rate; GFAP = Glial Fibrillary Acid Protein; MCI = Mild Cognitive Impairment; NH/OPI = Native Hawaiian or Other Pacific Islander; NfL = Neurofilament Light Chain.

**Table S2. Overall model fit statistics for independent regression models assessing associations between baseline plasma biomarkers and cognition**

| | | MODEL 1 | | | | MODEL 2 | | | |
|---|---|---|---|---|---|---|---|---|---|
| | | R2 | R2-ADJ | AIC | BIC | R2 | R2-ADJ | AIC | BIC |
| PACC5 | Aβ42/40 | .267 | .255 | 1601.766 | 1649.568 | .267 | .240 | 1146.737 | 1210.400 |
| | p-tau181 | .311 | .300 | 1574.586 | 1622.465 | .312 | .287 | 1128.415 | 1192.198 |
| | p-tau217 | .352 | .341 | 1393.219 | 1440.466 | .327 | .298 | 919.154 | 980.417 |
| | NfL | .277 | .266 | 1593.282 | 1641.084 | .287 | .260 | 1135.804 | 1199.467 |
| | GFAP | .318 | .307 | 1560.090 | 1607.892 | .312 | .287 | 1121.313 | 1184.975 |
| MEMORY | Aβ42/40 | .093 | .079 | 1804.664 | 1852.790 | .138 | .108 | 1286.656 | 1350.836 |
| | p-tau181 | .164 | .151 | 1764.256 | 1812.438 | .206 | .177 | 1260.704 | 1325.001 |
| | p-tau217 | .202 | .189 | 1639.227 | 1686.874 | .224 | .191 | 1043.633 | 1105.360 |
| | NfL | .120 | .107 | 1786.616 | 1834.741 | .185 | .156 | 1264.062 | 1328.242 |
| | GFAP | .172 | .159 | 1751.170 | 1799.296 | .209 | .181 | 1251.622 | 1315.802 |
| EXECUTIVE | Aβ42/40 | .061 | .047 | 1625.498 | 1673.624 | .067 | .034 | 1172.824 | 1237.004 |
| | p-tau181 | .099 | .085 | 1608.402 | 1656.583 | .107 | .076 | 1161.704 | 1226.001 |
| | p-tau217 | .089 | .075 | 1488.584 | 1536.230 | .084 | .046 | 962.023 | 1023.750 |
| | NfL | .076 | .061 | 1616.344 | 1664.470 | .096 | .064 | 1160.149 | 1224.330 |
| | GFAP | .081 | .066 | 1613.361 | 1661.486 | .099 | .067 | 1158.683 | 1222.863 |
| ATTENTION | Aβ42/40 | .027 | .011 | 1771.584 | 1819.709 | .044 | .010 | 1247.021 | 1311.202 |
| | p-tau181 | .062 | .048 | 1757.467 | 1805.648 | .082 | .050 | 1236.775 | 1301.073 |
| | p-tau217 | .044 | .029 | 1610.146 | 1657.793 | .066 | .027 | 984.774 | 1046.501 |
| | NfL | .047 | .032 | 1759.340 | 1807.465 | .069 | .036 | 1236.108 | 1300.288 |
| | GFAP | .048 | .033 | 1758.506 | 1806.632 | .066 | .032 | 1237.507 | 1301.687 |
| LANGUAGE | Aβ42/40 | .092 | .078 | 1497.418 | 1545.543 | .106 | .074 | 1058.778 | 1122.958 |
| | p-tau181 | .133 | .119 | 1474.625 | 1522.807 | .152 | .122 | 1044.162 | 1108.460 |
| | p-tau217 | .137 | .123 | 1390.590 | 1438.237 | .139 | .103 | 893.733 | 955.460 |
| | NfL | .108 | .094 | 1486.960 | 1535.085 | .135 | .104 | 1045.374 | 1109.555 |
| | GFAP | .129 | .116 | 1472.838 | 1520.963 | .149 | .119 | 1038.557 | 1102.737 |
| VISUOSPATIAL | Aβ42/40 | .006 | -.009 | 2101.074 | 2149.199 | .041 | .007 | 1501.611 | 1565.791 |
| | p-tau181 | .035 | .020 | 2091.118 | 2139.299 | .089 | .056 | 1488.645 | 1552.942 |
| | p-tau217 | .027 | .011 | 1771.227 | 1818.854 | .092 | .054 | 1029.509 | 1091.236 |
| | NfL | .020 | .004 | 2093.199 | 2141.324 | .065 | .032 | 1491.287 | 1555.468 |
| | GFAP | .020 | .005 | 2092.699 | 2140.824 | .055 | .021 | 1495.793 | 1559.973 |
| PHONEMIC | Aβ42/40 | .026 | .010 | 1657.239 | 1705.326 | .076 | .043 | 1149.288 | 1213.390 |
| | p-tau181 | .030 | .015 | 1661.339 | 1709.483 | .088 | .056 | 1153.173 | 1217.393 |
| | p-tau217 | .032 | .016 | 1621.917 | 1669.544 | .090 | .052 | 1007.692 | 1069.374 |
| | NfL | .030 | .015 | 1654.711 | 1702.799 | .091 | .058 | 1142.627 | 1206.728 |
| | GFAP | .027 | .012 | 1656.144 | 1704.232 | .084 | .051 | 1145.596 | 1209.697 |

***Note:*** Linear regression models assessing associations between plasma biomarkers (independently) and cognition at baseline. Plasma biomarkers have been log-transformed (except Aβ42/40) and scaled (centered and z-transformed) to permit direct comparisons. Model 1 adjusted for age, sex, race, ethnicity, education, and *APOE*. Model 2 additional adjusted for eGFR, BMI, diabetes, and hypertension. Variance explained is not directly comparable across models due to missing covariate data. ***Abbreviations:*** AIC = Akaike Information Criterion; ADJ = Adjusted; BIC = Bayesian Information Criterion; PACC5 = Preclinical Alzheimer's Cognitive Composite; R2 (R-squared) = Coefficient of Determination.

**Table S3. Simultaneous linear regression models evaluating associations between baseline plasma biomarkers and baseline cognition (all markers in the models simultaneously)**

| | | MODEL 1 | | | | MODEL 2 | | | | |
|---|---|---|---|---|---|---|---|---|---|---|
| | | **Est** | **CI** | ***p*** | **R2/R2-ADJ** | | **Est** | **CI** | ***p*** | **R2/R2-ADJ** |
| **PACC5** | **Aβ42/40** | 0.079 | 0.002 – 0.155 | .044* | .374 / .357 | **Aβ42/40** | 0.056 | -0.044 – 0.155 | .270 | .359 / .322 |
| | **p-tau181** | 0.089 | -0.028 – 0.205 | .134 | | **p-tau181** | 0.093 | -0.064 – 0.251 | .244 | |
| | **p-tau217** | **-0.390** | **-0.529 – -0.250** | **<.001** | | **p-tau217** | -0.384 | -0.567 – -0.202 | **<.001** | |
| | **NfL** | -0.002 | -0.122 – 0.117 | .972 | | **NfL** | -0.095 | -0.245 – 0.055 | .215 | |
| | **GFAP** | -0.148 | -0.261 – -0.035 | .010* | | **GFAP** | -0.133 | -0.276 – 0.010 | .068 | |
| **MEMORY** | **Aβ42/40** | 0.073 | -0.011 – 0.157 | .090 | .217 / .196 | **Aβ42/40** | 0.021 | -0.082 – 0.124 | .683 | .255 / .213 |
| | **p-tau181** | 0.070 | -0.058 – 0.199 | .284 | | **p-tau181** | 0.104 | -0.063 – 0.271 | .222 | |
| | **p-tau217** | **-0.396** | **-0.548 – -0.243** | **<.001** | | **p-tau217** | **-0.438** | **-0.633 – -0.242** | **<.001** | |
| | **NfL** | -0.008 | -0.147 – 0.130 | .910 | | **NfL** | -0.131 | -0.308 – 0.045 | .144 | |
| | **GFAP** | **-0.197** | **-0.330 – -0.063** | **.004** | | **GFAP** | -0.176 | -0.343 – -0.008 | .040* | |
| **EXECUTIVE** | **Aβ42/40** | 0.085 | -0.005 – 0.176 | .065 | .104 / .080 | **Aβ42/40** | 0.066 | -0.052 – 0.184 | .272 | .106 / .055 |
| | **p-tau181** | 0.000 | -0.123 – 0.124 | .995 | | **p-tau181** | -0.009 | -0.170 – 0.151 | .908 | |
| | **p-tau217** | -0.185 | -0.337 – -0.032 | .018* | | **p-tau217** | -0.110 | -0.292 – 0.073 | .238 | |
| | **NfL** | -0.028 | -0.145 – 0.089 | .638 | | **NfL** | -0.067 | -0.216 – 0.082 | .375 | |
| | **GFAP** | -0.073 | -0.190 – 0.044 | .222 | | **GFAP** | -0.138 | -0.290 – 0.014 | .076 | |
| **ATTENTION** | **Aβ42/40** | -0.013 | -0.091 – 0.066 | .749 | .062 / .037 | **Aβ42/40** | -0.045 | -0.138 – 0.048 | .345 | .089 / .037 |
| | **p-tau181** | -0.048 | -0.164 – 0.067 | .411 | | **p-tau181** | -0.092 | -0.236 – 0.052 | .210 | |
| | **p-tau217** | -0.120 | -0.278 – 0.039 | .139 | | **p-tau217** | -0.058 | -0.230 – 0.114 | .508 | |
| | **NfL** | -0.081 | -0.194 – 0.032 | .159 | | **NfL** | -0.100 | -0.236 – 0.037 | .151 | |
| | **GFAP** | -0.054 | -0.174 – 0.066 | .374 | | **GFAP** | -0.087 | -0.227 – 0.053 | .223 | |
| **LANGUAGE** | **Aβ42/40** | 0.022 | -0.046 – 0.091 | .520 | .141 / .118 | **Aβ42/40** | -0.025 | -0.114 – 0.064 | .575 | .174 / .127 |
| | **p-tau181** | -0.017 | -0.133 – 0.098 | .769 | | **p-tau181** | -0.012 | -0.167 – 0.143 | .882 | |
| | **p-tau217** | **-0.182** | **-0.308 – -0.056** | **.005** | | **p-tau217** | -0.192 | -0.336 – -0.048 | .009* | |
| | **NfL** | -0.011 | -0.131 – 0.109 | .855 | | **NfL** | -0.085 | -0.230 – 0.059 | .245 | |
| | **GFAP** | -0.085 | -0.195 – 0.024 | .127 | | **GFAP** | -0.108 | -0.245 – 0.029 | .121 | |
| **VISUOSPATIAL** | **Aβ42/40** | -0.039 | -0.135 – 0.057 | .423 | .036 / .010 | **Aβ42/40** | -0.085 | -0.199 – 0.029 | .145 | .113 / .062 |
| | **p-tau181** | 0.042 | -0.086 – 0.169 | .520 | | **p-tau181** | -0.035 | -0.185 – 0.115 | .643 | |
| | **p-tau217** | -0.181 | -0.340 – -0.022 | .026* | | **p-tau217** | -0.200 | -0.377 – -0.022 | .027* | |
| | **NfL** | -0.099 | -0.279 – 0.081 | .279 | | **NfL** | -0.186 | -0.383 – 0.011 | .063 | |
| | **GFAP** | 0.022 | -0.127 – 0.172 | .769 | | **GFAP** | 0.089 | -0.079 – 0.258 | .296 | |
| **PHONEMIC** | **Aβ42/40** | -0.011 | -0.117 – 0.094 | .833 | .033 / .007 | **Aβ42/40** | -0.050 | -0.180 – 0.080 | .450 | .106 / .055 |
| | **p-tau181** | -0.041 | -0.185 – 0.102 | .571 | | **p-tau181** | -0.058 | -0.243 – 0.128 | .540 | |
| | **p-tau217** | 0.014 | -0.141 – 0.170 | .855 | | **p-tau217** | 0.010 | -0.183 – 0.204 | .918 | |
| | **NfL** | -0.019 | -0.142 – 0.103 | .758 | | **NfL** | -0.098 | -0.249 – 0.053 | .203 | |
| | **GFAP** | -0.014 | -0.134 – 0.106 | .817 | | **GFAP** | -0.056 | -0.205 – 0.093 | .461 | |

***Note:*** Simultaneous linear regression models assessing associations between baseline plasma biomarkers (entered simultaneously as predictors) and cognition at baseline. Plasma biomarkers were log-transformed (except Aβ42/40) and scaled (centered and z-transformed) to permit direct comparisons. Significant effects bolded ($p < .05$). * = no longer significant after FDR. ***Abbreviations:*** Aβ = Amyloid Beta; CI = 95% Confidence Interval; GFAP = Glial Fibrillary Acidic Protein; NfL = Neurofilament Light Chain; PACC5 = Preclinical Alzheimer's Cognitive Composite; p-tau = Phosphorylated Tau; R2 (R-squared) = Coefficient of Determination.

**Table S4. Overall model fit statistics for independent linear mixed effect model results assessing associations between baseline plasma biomarkers and change in cognition**

| | | MODEL 1 | | | | | MODEL 2 | | | | | |
|---|---|---|---|---|---|---|---|---|---|---|---|---|
| | | MR2 | CR2 | $\sigma^2$ | τ00 | ICC | | MR2 | CR2 | $\sigma^2$ | τ00 | ICC |
| PACC5 | **Aβ42/40** | .27 | .91 | 0.14 | 0.94 | 0.87 | **Aβ42/40** | .28 | .91 | 0.14 | 1.00 | 0.88 |
| | **p-tau181** | .31 | .91 | 0.14 | 0.88 | 0.87 | **p-tau181** | .31 | .91 | 0.14 | 0.93 | 0.87 |
| | **p-tau217** | .34 | .90 | 0.14 | 0.76 | 0.85 | **p-tau217** | .32 | .90 | 0.14 | 0.85 | 0.86 |
| | **NfL** | .28 | .91 | 0.14 | 0.92 | 0.87 | **NfL** | .29 | .91 | 0.14 | 0.97 | 0.88 |
| | **GFAP** | .31 | .91 | 0.14 | 0.88 | 0.86 | **GFAP** | .32 | .91 | 0.14 | 0.94 | 0.87 |
| MEMORY | **Aβ42/40** | .12 | .83 | 0.27 | 1.14 | 0.81 | **Aβ42/40** | .16 | .84 | 0.27 | 1.19 | 0.82 |
| | **p-tau181** | .18 | .83 | 0.27 | 1.03 | 0.80 | **p-tau181** | .21 | .84 | 0.27 | 1.09 | 0.80 |
| | **p-tau217** | .22 | .83 | 0.27 | 0.93 | 0.78 | **p-tau217** | .23 | .84 | 0.28 | 1.01 | 0.79 |
| | **NfL** | .13 | .83 | 0.27 | 1.11 | 0.81 | **NfL** | .18 | .84 | 0.27 | 1.14 | 0.81 |
| | **GFAP** | .18 | .83 | 0.27 | 1.03 | 0.80 | **GFAP** | .21 | .84 | 0.27 | 1.09 | 0.80 |
| EXECUTIVE | **Aβ42/40** | .07 | .74 | 0.29 | 0.73 | 0.72 | **Aβ42/40** | .08 | .75 | 0.29 | 0.78 | 0.73 |
| | **p-tau181** | .11 | .74 | 0.29 | 0.68 | 0.70 | **p-tau181** | .12 | .75 | 0.29 | 0.73 | 0.72 |
| | **p-tau217** | .12 | .72 | 0.29 | 0.63 | 0.69 | **p-tau217** | .12 | .74 | 0.30 | 0.69 | 0.70 |
| | **NfL** | .08 | .74 | 0.29 | 0.71 | 0.71 | **NfL** | .10 | .75 | 0.29 | 0.75 | 0.72 |
| | **GFAP** | .09 | .74 | 0.29 | 0.70 | 0.71 | **GFAP** | .11 | .75 | 0.29 | 0.74 | 0.72 |
| ATTENTION | **Aβ42/40** | .04 | .74 | 0.35 | 0.95 | 0.73 | **Aβ42/40** | .06 | .79 | 0.29 | 0.98 | 0.77 |
| | **p-tau181** | .07 | .74 | 0.35 | 0.90 | 0.72 | **p-tau181** | .08 | .78 | 0.29 | 0.93 | 0.76 |
| | **p-tau217** | .07 | .70 | 0.35 | 0.74 | 0.68 | **p-tau217** | .09 | .75 | 0.27 | 0.72 | 0.73 |
| | **NfL** | .06 | .74 | 0.35 | 0.92 | 0.72 | **NfL** | .07 | .78 | 0.29 | 0.95 | 0.77 |
| | **GFAP** | .06 | .74 | 0.35 | 0.92 | 0.72 | **GFAP** | .07 | .78 | 0.29 | 0.95 | 0.77 |
| LANGUAGE | **Aβ42/40** | .10 | .66 | 0.36 | 0.59 | 0.62 | **Aβ42/40** | .11 | .63 | 0.44 | 0.62 | 0.59 |
| | **p-tau181** | .13 | .66 | 0.36 | 0.56 | 0.61 | **p-tau181** | .13 | .63 | 0.44 | 0.58 | 0.57 |
| | **p-tau217** | .17 | .83 | 0.16 | 0.61 | 0.80 | **p-tau217** | .17 | .85 | 0.15 | 0.69 | 0.82 |
| | **NfL** | .11 | .66 | 0.36 | 0.58 | 0.62 | **NfL** | .12 | .63 | 0.44 | 0.59 | 0.57 |
| | **GFAP** | .13 | .66 | 0.36 | 0.57 | 0.61 | **GFAP** | .14 | .63 | 0.44 | 0.58 | 0.57 |
| VISUO-SPATIAL | **Aβ42/40** | .01 | .63 | 0.70 | 1.16 | 0.62 | **Aβ42/40** | .03 | .66 | 0.65 | 1.18 | 0.64 |
| | **p-tau181** | .04 | .63 | 0.70 | 1.10 | 0.61 | **p-tau181** | .07 | .65 | 0.65 | 1.09 | 0.63 |
| | **p-tau217** | .03 | .52 | 0.69 | 0.69 | 0.50 | **p-tau217** | .06 | .46 | 0.63 | 0.46 | 0.42 |
| | **NfL** | .02 | .63 | 0.70 | 1.14 | 0.62 | **NfL** | .05 | .65 | 0.65 | 1.13 | 0.64 |
| | **GFAP** | .03 | .63 | 0.70 | 1.13 | 0.62 | **GFAP** | .04 | .66 | 0.65 | 1.16 | 0.64 |
| PHONEMIC | **Aβ42/40** | .04 | .57 | 0.61 | 0.75 | 0.55 | **Aβ42/40** | .07 | .61 | 0.52 | 0.73 | 0.58 |
| | **p-tau181** | .04 | .57 | 0.61 | 0.74 | 0.55 | **p-tau181** | .08 | .61 | 0.52 | 0.72 | 0.58 |
| | **p-tau217** | .05 | .67 | 0.43 | 0.81 | 0.65 | **p-tau217** | .09 | .78 | 0.26 | 0.81 | 0.76 |
| | **NfL** | .04 | .57 | 0.61 | 0.74 | 0.55 | **NfL** | .08 | .61 | 0.52 | 0.71 | 0.58 |
| | **GFAP** | .04 | .57 | 0.61 | 0.74 | 0.55 | **GFAP** | .08 | .61 | 0.52 | 0.71 | 0.58 |

***Note:*** Longitudinal mixed effect models assessing associations between plasma biomarkers (independent) and cognition. P-tau217 generally showed stronger associations with cognitive change based on fixed-effect variance (marginal $R^2$), overall model performance and variability in cognitive outcomes were comparable however across biomarkers and adjustment levels. ***Abbreviations*:** Aβ = Amyloid Beta; CR2 = Conditional (Fixed & Random) R-Squared; GFAP = Glial Fibrillary Acidic Protein; ICC = Intraclass Correlation; MR2 = Marginal (Fixed Effects) R-Squared; NfL = Neurofilament Light Chain; PACC5 = Preclinical Alzheimer's Cognitive Composite; p-tau = Phosphorylated Tau; $\sigma^2$= Sigma Squared (Residual Variance); $\tau_{00}$ = *tau-squared* (Random Intercept).

**Table S5. Simultaneous longitudinal linear mixed effect model results and fit statistics**

| | | MODEL 1 | | | | | | | MODEL 2 | | | | | | |
|---|---|---|---|---|---|---|---|---|---|---|---|---|---|---|---|
| | | Est | CI | p | MR2/ CR2 | $\sigma^2$ | τ00 | ICC | Est | CI | p | MR2/ CR2 | $\sigma^2$ | τ00 | ICC |
| PACC5 | **Aβ42/40** | 0.09 | 0.01 – 0.17 | .040* | .356/.899 | 0.1 | 0.7 | 0.8 | 0.09 | -0.02 – 0.20 | .130 | .358/.906 | 0.1 | 0.8 | 0.9 |
| | p-tau181 | 0.09 | -0.02 – 0.20 | .120 | | | | | 0.10 | -0.06 – 0.25 | .230 | | | | |
| | **p-tau217** | **-0.40** | **-0.52 – -0.23** | **<.001** | | | | | **-0.37** | **-0.55 – -0.18** | **<.001** | | | | |
| | NfL | -0.02 | -0.14 – 0.10 | .740 | | | | | -0.10 | -0.25 – 0.04 | .170 | | | | |
| | **GFAP** | -0.10 | -0.25 – -0.02 | .020* | | | | | -0.13 | -0.27 – 0.01 | .070 | | | | |
| MEMORY | **Aβ42/40** | 0.11 | 0.02 – 0.20 | .010* | .238/.822 | 0.3 | 0.9 | 0.8 | 0.11 | -0.01 – 0.22 | .070 | .266/.836 | 0.3 | 1 | 0.8 |
| | p-tau181 | 0.08 | -0.04 – 0.20 | .210 | | | | | 0.09 | -0.07 – 0.25 | .270 | | | | |
| | **p-tau217** | **-0.50** | **-0.60 – -0.30** | **<.001** | | | | | **-0.45** | **-0.65 – -0.26** | **<.001** | | | | |
| | NfL | -0.02 | -0.15 – 0.11 | .800 | | | | | -0.12 | -0.29 – 0.05 | .190 | | | | |
| | **GFAP** | **-0.19** | **-0.31 – -0.06** | **.003** | | | | | -0.17 | -0.33 – -0.02 | .030* | | | | |
| EXECUTIVE | Aβ42/40 | 0.05 | -0.03 – 0.13 | .200 | .129/.728 | 0.3 | 0.6 | 0.7 | 0.04 | -0.07 – 0.14 | .460 | .134/.734 | 0.3 | 0.7 | 0.7 |
| | p-tau181 | 0.02 | -0.09 – 0.12 | .750 | | | | | 0.02 | -0.12 – 0.16 | .780 | | | | |
| | **p-tau217** | **-0.30** | **-0.40 – -0.14** | **<.001** | | | | | -0.22 | -0.38 – -0.06 | .008* | | | | |
| | NfL | -0.02 | -0.12 – 0.08 | .700 | | | | | -0.07 | -0.20 – 0.07 | .330 | | | | |
| | GFAP | -0.10 | -0.18 – 0.04 | .190 | | | | | -0.12 | -0.26 – 0.02 | .090 | | | | |
| ATTENTION | Aβ42/40 | -0.02 | -0.09 – 0.05 | .580 | .083/.705 | 0.3 | 0.7 | 0.7 | -0.01 | -0.11 – 0.08 | .760 | .092/.755 | 0.3 | 0.7 | 0.7 |
| | p-tau181 | -0.01 | -0.11 – 0.09 | .810 | | | | | -0.02 | -0.15 – 0.11 | .730 | | | | |
| | **p-tau217** | **-0.22** | **-0.37 – -0.07** | **.005** | | | | | -0.15 | -0.31 – 0.01 | .070 | | | | |
| | NfL | -0.07 | -0.16 – 0.02 | .140 | | | | | -0.09 | -0.20 – 0.02 | .120 | | | | |
| | GFAP | -0.05 | -0.16 – 0.05 | .330 | | | | | -0.08 | -0.20 – 0.04 | .210 | | | | |
| LANGUAGE | Aβ42/40 | 0.05 | -0.02 – 0.12 | .160 | .171/.829 | 0.2 | 0.6 | 0.8 | 0.01 | -0.08 – 0.11 | .790 | .200/.853 | 0.2 | 0.7 | 0.8 |
| | p-tau181 | 0.00 | -0.10 – 0.10 | .960 | | | | | -0.01 | -0.15 – 0.13 | .870 | | | | |
| | **p-tau217** | **-0.24** | **-0.36 – -0.11** | **<.001** | | | | | **-0.26** | **-0.41 – -0.10** | **.002** | | | | |
| | NfL | -0.04 | -0.15 – 0.07 | .500 | | | | | -0.10 | -0.24 – 0.04 | .160 | | | | |
| | GFAP | -0.07 | -0.17 – 0.03 | .190 | | | | | -0.09 | -0.21 – 0.04 | .190 | | | | |
| VISUO-SPATIAL | Aβ42/40 | 0.02 | -0.06 – 0.10 | .550 | .042/.500 | 0.7 | 0.6 | 0.5 | 0.03 | -0.06 – 0.12 | .520 | .071/.458 | 0.6 | 0.5 | 0.4 |
| | p-tau181 | 0.00 | -0.09 – 0.10 | .960 | | | | | -0.03 | -0.15 – 0.09 | .610 | | | | |
| | p-tau217 | -0.13 | -0.27 – 0.00 | .060 | | | | | -0.13 | -0.26 – -0.00 | .050 | | | | |
| | NfL | -0.08 | -0.21 – 0.06 | .270 | | | | | -0.16 | -0.31 – -0.01 | .050 | | | | |
| | GFAP | -0.03 | -0.15 – 0.08 | .580 | | | | | 0.05 | -0.08 – 0.18 | .430 | | | | |
| PHONEMIC | Aβ42/40 | 0.00 | -0.10 – 0.10 | .950 | .049/.664 | 0.4 | 0.8 | 0.7 | -0.04 | -0.17 – 0.09 | .560 | .102/.779 | 0.3 | 0.8 | 0.8 |
| | p-tau181 | -0.02 | -0.15 – 0.11 | .750 | | | | | -0.06 | -0.23 – 0.11 | .520 | | | | |
| | p-tau217 | -0.07 | -0.22 – 0.08 | .380 | | | | | -0.08 | -0.26 – 0.11 | .410 | | | | |
| | NfL | -0.03 | -0.14 – 0.08 | .640 | | | | | -0.08 | -0.22 – 0.07 | .310 | | | | |
| | GFAP | 0.00 | -0.12 – 0.11 | .980 | | | | | -0.06 | -0.20 – 0.08 | .410 | | | | |

***Note:*** Simultaneous longitudinal mixed effects models (baseline plasma biomarkers entered simultaneously) assessing biomarker–cognition associations. Simultaneous modeling revealed that p-tau217 consistently accounted for the largest independent association with cognition across domains, while associations for other biomarkers were substantially attenuated compared with independent models. Plasma biomarkers were log-transformed (except Aβ42/40) and scaled (centered and z-transformed). Significant coefficients bolded ($p <$ .05). * = no longer significant after FDR. ***Abbreviations***: CI = 95% Confidence Interval; MR2 = Marginal (Fixed Effects) R-Squared; CR2 = Conditional (Fixed & Random) R-Squared; $\sigma^2$= Sigma Squared (Residual Variance); $\tau_{00}$ = *tau-squared (Random Intercept);* PACC5 = Preclinical Alzheimer's Cognitive Composite; Aβ

= Amyloid Beta; p-tau = Phosphorylated Tau; NfL = Neurofilament Light Chain; GFAP = Glial Fibrillary Acidic Protein

**Table S6. Independent and simultaneous linear mixed effect models assessing effect modification by cognitive status on plasma biomarker-cognition associations**

| | | INDEPENDENT MODELS | | | JOINT MODELS | | |
|---|---|---|---|---|---|---|---|
| | | CU slope (SE) | MCI slope (SE) | *p*-INT | CU slope (SE) | MCI slope (SE) | *p*-INT |
| PACC5 | Aβ42/40 | **0.02 (0.04)** | **0.18 (0.07)** | **.011** | 0.04 (0.04) | 0.14 (0.09) | .193 |
| | p-tau181 | **-0.00 (0.04)** | **-0.14 (0.08)** | **.048** | 0.05 (0.06) | -0.04 (0.13) | .451 |
| | p-tau217 | **-0.01 (0.05)** | **-0.16 (0.09)** | **.038** | -0.01 (0.06) | -0.07 (0.13) | .626 |
| | NfL | 0.00 (0.04) | -0.04 (0.07) | .431 | -0.04 (0.06) | 0.05 (0.10) | .264 |
| | GFAP | **0.01 (0.04)** | **-0.15 (0.07)** | **.008** | 0.01 (0.05) | -0.09 (0.10) | .290 |
| MEMORY | Aβ42/40 | **-0.01 (0.05)** | **0.26 (0.09)** | **<.001** | 0.02 (0.04) | 0.19 (0.10) | .052 |
| | p-tau181 | **-0.04 (0.05)** | **-0.20 (0.09)** | **.045** | 0.04 (0.06) | -0.05 (0.14) | .491 |
| | p-tau217 | **-0.04 (0.06)** | **-0.27 (0.10)** | **.010** | -0.02 (0.06) | -0.09 (0.14) | .563 |
| | NfL | -0.02 (0.05) | -0.07 (0.09) | .537 | -0.07 (0.06) | 0.11 (0.11) | .050 |
| | GFAP | **-0.01 (0.05)** | **-0.27 (0.08)** | **<.001** | **0.02 (0.06)** | **-0.21 (0.11)** | **.019** |
| EXECUTIVE | Aβ42/40 | 0.09 (0.04) | 0.03 (0.09) | .436 | 0.06 (0.05) | -0.04 (0.11) | .325 |
| | p-tau181 | -0.06 (0.06) | -0.12 (0.10) | .515 | -0.01 (0.07) | -0.11 (0.14) | .401 |
| | p-tau217 | -0.07 (0.06) | -0.10 (0.12) | .757 | -0.01 (0.07) | 0.01 (0.16) | .862 |
| | NfL | -0.05 (0.06) | -0.01 (0.09) | .624 | -0.05 (0.07) | 0.14 (0.12) | .053 |
| | GFAP | -0.04 (0.06) | -0.10 (0.10) | .514 | -0.02 (0.07) | -0.16 (0.15) | .288 |
| ATTENTION | Aβ42/40 | 0.01 (0.04) | 0.05 (0.07) | .492 | -0.05 (0.04) | -0.00 (0.09) | .570 |
| | p-tau181 | -0.04 (0.05) | -0.10 (0.09) | .514 | -0.03 (0.06) | -0.14 (0.14) | .417 |
| | p-tau217 | -0.05 (0.06) | -0.12 (0.10) | .435 | -0.01 (0.06) | 0.04 (0.14) | .619 |
| | NfL | -0.04 (0.06) | -0.07 (0.09) | .637 | -0.01 (0.06) | -0.02 (0.10) | .911 |
| | GFAP | -0.03 (0.05) | -0.09 (0.09) | .413 | -0.03 (0.06) | -0.06 (0.11) | .734 |
| LANGUAGE | Aβ42/40 | **-0.03 (0.04)** | **0.11 (0.07)** | **.022** | -0.03 (0.05) | 0.03 (0.09) | .473 |
| | p-tau181 | -0.06 (0.04) | -0.12 (0.08) | .478 | -0.11 (0.08) | -0.00 (0.15) | .407 |
| | p-tau217 | -0.01 (0.05) | -0.15 (0.09) | .059 | 0.04 (0.08) | -0.09 (0.14) | .266 |
| | NfL | -0.00 (0.05) | -0.07 (0.07) | .224 | 0.01 (0.07) | 0.00 (0.11) | .940 |
| | GFAP | -0.02 (0.04) | -0.13 (0.07) | .060 | -0.00 (0.06) | -0.06 (0.11) | .519 |
| VISUO SPATIAL | Aβ42/40 | 0.05 (0.04) | 0.06 (0.09) | .953 | 0.05 (0.04) | 0.06 (0.11) | .919 |
| | p-tau181 | -0.04 (0.05) | -0.17 (0.11) | .188 | 0.03 (0.06) | -0.10 (0.14) | .309 |
| | p-tau217 | -0.04 (0.05) | -0.23 (0.12) | .070 | -0.03 (0.06) | -0.19 (0.14) | .236 |
| | NfL | -0.08 (0.06) | -0.11 (0.11) | .724 | -0.17 (0.06) | -0.08 (0.14) | .504 |
| | GFAP | 0.01 (0.04) | -0.04 (0.08) | .502 | 0.10 (0.05) | 0.18 (0.14) | .549 |
| PHONEMIC | Aβ42/40 | -0.04 (0.07) | -0.02 (0.11) | .766 | -0.09 (0.08) | -0.05 (0.14) | .772 |
| | p-tau181 | -0.08 (0.07) | 0.00 (0.11) | .342 | -0.13 (0.12) | 0.01 (0.20) | .404 |
| | p-tau217 | 0.02 (0.07) | 0.05 (0.11) | .734 | 0.11 (0.11) | 0.02 (0.19) | .576 |
| | NfL | -0.06 (0.07) | 0.02 (0.10) | .282 | -0.05 (0.10) | 0.07 (0.15) | .303 |
| | GFAP | -0.06 (0.06) | -0.01 (0.10) | .439 | -0.04 (0.08) | -0.07 (0.14) | .806 |

***Note***. Fully-adjusted independent and simultaneous longitudinal mixed effects models assessing whether biomarker–cognition associations are modified by cognitive status. Interaction effects represent the difference in biomarker-cognition slopes for MCI compared with CU. Across several cognitive domains, higher biomarker burden was associated with poorer cognitive performance, with these associations generally more pronounced among participants with mild cognitive impairment than among cognitively unimpaired participants, although patterns varied by biomarker and domain. Plasma biomarkers were log-transformed (except Aβ42/40) and scaled (centered and z-transformed) prior to analysis. Statistically significant coefficients are shown in bold (*p*-INT <

0.05). ***Abbreviations***: Aβ = amyloid-β; CU = Cognitively Unimpaired; GFAP = glial fibrillary acidic protein; MCI = Mild Cognitive Impairment; NfL = neurofilament light chain; PACC5 = Preclinical Alzheimer's Cognitive Composite; p-tau = phosphorylated tau; *p*-INT = interaction *p*-value; SE = Standard Error.
